# Change in movement disorder specialist attitudes to genetic testing after implementation of PD GENEration

**DOI:** 10.1101/2025.07.29.25332392

**Authors:** Jesse Wang, Vanessa Ibrahim, Roy N. Alcalay, Julian Agin-Liebes, Martha Nance, James C. Beck, Margaret E. Caulfield, Anna Naito, Kamalini Ghosh Galvelis, Anne-Marie Wills

## Abstract

**Purpose:** Despite advances in recent years, genetic testing for Parkinson’s disease (PD) is still underutilized in clinical practice. A 2019 questionnaire of movement disorder specialists found low rates of genetic testing in PD and barriers such as cost and insurance coverage. Since that time, PD GENEration as well as several international programs have broadly increased access to genetic testing and counseling for people with PD. A new survey sent out in 2024 examined how genetic testing for PD has changed over the past 5 years.

**Methods:** Between October 2024-January 2025, 621 movement disorders specialists from the Parkinson’s Study Group (PSG) were invited by email to complete a questionnaire assessing knowledge, attitudes, and barriers to genetic testing in PD based on the 2019 survey.

**Results:** 119 total PSG clinicians from the United States and Canada responded to the questionnaire. When compared to results from 2019, 2024 survey respondents reported increased comfort and willingness to order genetic testing, including reduced fears of negative repercussions. PD GENEration-affiliated sites reported significantly higher genetic testing volume and patients with known genetic variants.

**Conclusions:** 2024 survey respondents report greater comfort with genetic testing in PD compared to 2019, concurrent with the launch of PD GENEration.

## INTRODUCTION

The landscape of genetic testing in Parkinson’s disease is rapidly evolving^1^. In 2019, Alcalay et al. surveyed 178 movement disorders specialists and experienced clinical trialists who were part of the Parkinson Study Group (PSG), representing 131 sites in the United States and 15 in Canada to assess their knowledge and attitudes about genetic testing for PD^2^. They found that 41% of respondents had not referred any PD patients for genetic testing in the prior year, and 87% of respondents had ordered 10 or fewer genetic tests in the prior year. The main barriers reported by respondents were lack of insurance coverage for genetic testing and the impression that genetic results would not change therapy. This was despite the fact that a high percentage of people with Parkinson’s disease (PWP) are interested in receiving genetic testing and genetic counseling^3–5^.

Since the survey by Alcalay et al. was published, a number of events have occurred that have impacted access to Parkinson’s disease genetic testing. First, the Parkinson’s Foundation launched the PD GENEration study (ClinicalTrials.gov identifiers: NCT04057794 and NCT04994015) in August 2019^6–9^. The study, initially only in the US but later expanded to other countries, provides testing and return of results via neurologists or genetic counselors for the 7 most common PD-related genes by a CLIA-certified laboratory at no cost to participants, without reliance on patient insurance. In addition to this 7-gene panel, in 2024 testing was expanded to include whole genome sequencing and an additional panel of 21 genes was added to the protocol. In the 5 years since the launch of PD GENEration, over 25,000 individuals with Parkinson’s disease have enrolled into the study, referred by clinicians, participating PD GENEration clinical trial sites, and through online self-referral, all supported via online advertising and social media^10^. From this study, approximately 13% of participants were identified with a PD-relevant variant, 9% if ancestry, age of onset, or family history were excluded^7^. Second, the Rostock International Parkinson’s Disease Study (ROPAD) study was also launched in 2019 (ClinicalTrials.gov identifier: NCT03866603) and has enrolled 12,580 participants mostly in Europe but also in the US and Latin America^11,12^. In the ROPAD study, participants underwent an initial screen for *LRRK2* and *GBA1* variants, followed by sequencing of a panel of 50 genes with 14.5% of participants found to have a positive PD-relevant genetic test. Third, the number of precision medicine clinical trials increased from just one *GBA1*-PD targeted therapy trial active in the US in 2019 to five in 2024^*^.

We hypothesized that the increased access to genetic testing through PD GENEration and, to some extent, ROPAD would impact North American clinicians’ attitudes towards genetic testing in PD. We therefore re-surveyed a large group of movement disorders clinicians in North

America and compared their answers to the 2019 survey results. Because PD GENEration recruits participants from many PSG sites, we analyzed separately the answers of sites that were involved in PD GENEration from those that were not.

## MATERIALS AND METHODS

### Participants

The PSG (Parkinson’s Study Group) is the largest Parkinson’s-disease clinical trial site network in North America, comprising over 155 credentialed sites across the US and Canada. To be credentialed as a member of the PSG, investigators must spend a significant percentage of their practice treating PD and conducting PD related research. Most PSG investigators have had fellowship training in movement disorders.

The survey protocol was approved by the Advarra Institutional Review Board (IRB). Given the low-risk nature of the protocol, acceptance of the invitation and an electronic consent were accepted as a waiver for paper consent.

A total of 621 clinician members of the PSG were invited to take part in an anonymous online survey (see *Supplementary Material*). Invitations to the anonymous online survey was sent three times between 10/15/24 and 1/6/25. Participants who chose to include their email contact information were provided with a $25 Amazon gift certificate.

### Questionnaire

Questions from the 2024 survey fell into the following categories:

1. Basic demographics of the participants (gender, age, location)
2. Questions about the clinician and their patient populations
3. Questions regarding current practice, accessibility, and barriers of genetic counseling (including involvement with PD GENEration)
4. General knowledge questions concerning PD genetics, including two case scenarios focused on *GBA1* and *LRRK2*-related PD

The questionnaire repeated many of the questions that were sent in the survey in 2019. Several new questions were added to assess PD GENEration involvement and its effectiveness in improving comfort with and access to genetic testing in Parkinson’s disease. The full questionnaire can be viewed in the Supplementary Material.

### Statistical Analysis

Because many of the surveyed respondents were affiliated with clinical sites involved in PD GENEration, data from PD GENEration and non-PD GENEration sites were analyzed separately. PD GENEration affiliates were defined as sites who reported that they were either involved as sites in PD GENEration or referred participants to the online enrollment center (“referral sites”). Because very few PSG sites were involved in the ROPAD study, sites were not separately analyzed. The original data from the 2019 survey was available and used to compare to the 2024 survey results. Descriptive statistics (percentages, means, and SD) were used to visually compare the responses across groups, and statistical significance between groups were calculated either by ANOVA, Kruskal-Wallis, or chi-square analysis. All statistical analyses were conducted using R [Version 4.4.2, 2024-10-31].

## RESULTS

Of the 621 total clinicians invited to take part in the anonymous online survey, only 119 participants responded and 113 completed the full questionnaire, which was significantly lower than the 178 PSG investigators who responded in 2019. Only twenty-two participants stated that they recalled responding to the 2019 survey. Because both surveys were anonymous, we were unable to match responses between the two.

Of the 119 responders, 108 were movement disorder specialists, 106 were PSG members, 74 reported practicing in an academic medical center, 67 were active in clinical trials for Parkinson’s disease, and 83 were involved with PD GENEration either as recruiting sites (58) or referral sites (25). Only 11 PSG sites were involved in the ROPAD study. Of the participants who chose to share their demographic data, there were 56 male respondents and 56 female respondents. All clinicians surveyed practice in the US or Canada (9) and reported working in clinics with a median PD population of 300 patients, (ranging from 4 to 2000).

### Current genetic testing practice

The distribution of responses between survey respondents (separated into groups by the 2019 respondents, non-PD GENE affiliated groups, and PD-GENE recruiting or referral sites) is shown below in **Figure 1**. First, the number of clinicians ordering more than 10 clinical tests per year (billed to patient insurance) did not significantly increase (12.3% in 2019 vs. 18.1% of PD GENEration respondents in 2024 vs 26.7% of non-PD GENEration respondents). By contrast, participants reporting more than 10 referrals/year to research testing that returns results to participants (e.g., PD GENEration, ROPAD, the Michael J. Fox Foundation Parkinson’s Progression Markers Initiative [PPMI]) increased from 15.5% in 2019 to 23.3% for non-PD GENEration sites and 60.2% for PD GENEration sites (Kruskal-Wallis p= 1.91 e-17).

**Figure 1:**
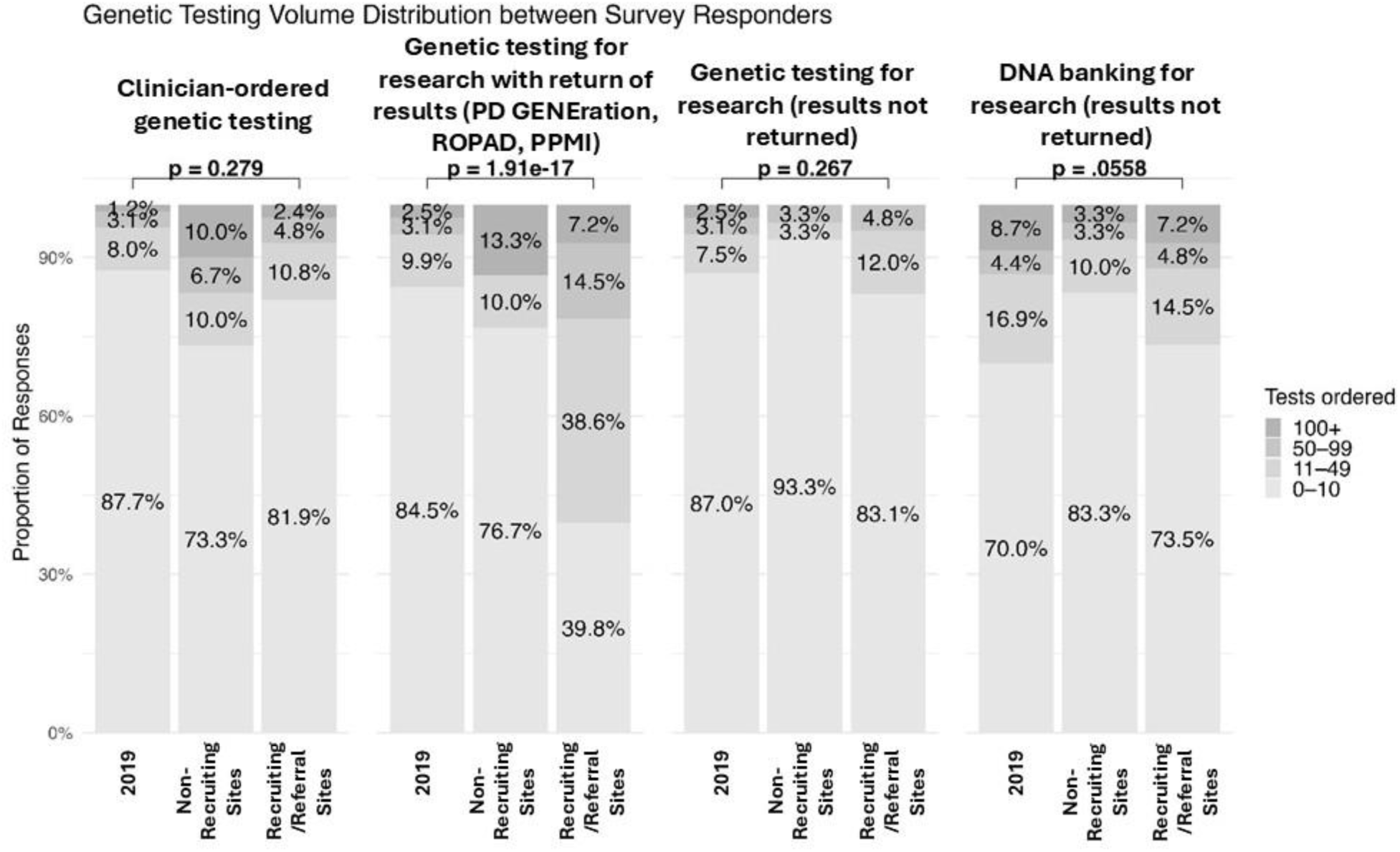
Number of reported genetic tests sent by respondents in the prior year: clinician-ordered genetic testing (clinical genetic tests billed to patient insurance), genetic testing where the participants receive their results (PD GENEration, ROPAD or PPMI), genetic testing for research that does not return results, and DNA banking for research. 2024 respondents are divided by sites not affiliated with PD GENEration (Non-Recruiting) and sites that are PD GENEration recruiting or referral sites (Recruiting/Referral). P-values for statistical significance across the three groups determined by Kruskal-Wallis test.

Genetic testing for research only (without return of results), and DNA banking did not significantly change between 2019 and 2024. Respondents reporting more than ten patients undergoing direct-to-consumer testing (e.g., 23andMe) remained stable at 20.0% for non-PD GENEration and 10.8% for PD GENEration sites compared to 14.6% in 2019 (data not shown).

In 2024, survey respondents also reported that patient interest and response to genetic testing has been largely positive, with only 16.8% of clinicians reporting more than 10 patients who refused genetic testing in the prior year across all sites.

As access to genetic testing has improved overall, the presence of known pathogenic variants has also increased among survey respondents. The number of clinicians reporting that they provided care to more than 10 patients with known genetic variants increased from 5.5% for *LRRK2* and 4.9% for *GBA1* in 2019, to 12.6% and 18.4%, respectively. The total number of *LRRK2* gene heterozygotes reported remained the same at 490 (although the 2024 survey included fewer sites) and *GBA1* heterozygotes increased from 402 in 2019 to 674 in 2024. Providing care for patients with known *PRKN*, *PINK1*, *SNCA*, *PARK7* and *VPS35* pathogenic variants has also become more prevalent. While only 2% of clinicians previously reported having more than two patients with these pathogenic variants, this increased to 21.8% of respondents in 2024, with 235 total cases reported. Other rare variants reported included *TWNK* and *ATP13A2* among others (9 total with 2 or fewer reported for each).

### Access to genetic counseling

In a situation where genetic testing would be ordered, a similar proportion (59.3%, compared to 61.2% in 2019) reported they would either involve a genetic counselor or recommend a genetic counseling referral prior to genetic testing for PD. A similar 20.3% of respondents (compared to 21.9% in 2019) reported they would provide the genetic counseling themselves. A majority (76%) reported having access to services of a genetic counselor within their institution or practice, largely unchanged from the 2019 survey (70%). A non-significant increase of 9.2% reported using genetic counseling via telephone or telemedicine compared to 4.5% in 2019.

### Obstacles to genetic testing

Respondents were then asked to grade the same obstacles from the 2019 survey that prevented them from ordering genetic tests on a scale from 0-100 (**Figure 2**). The most prevalent obstacles remained lack of insurance coverage/cost and the opinion that test results would not change their approach to patient care. However, all of the perceived barriers were significantly lower in 2024 compared to 2019 in both PD GENEration recruiting/referral and non-affiliated sites, including insurance barriers and concerns about repercussions. There was no significant difference in the perceived barriers between PD GENEration recruiting/referral sites and non-affiliated sites.

**Figure 2:**
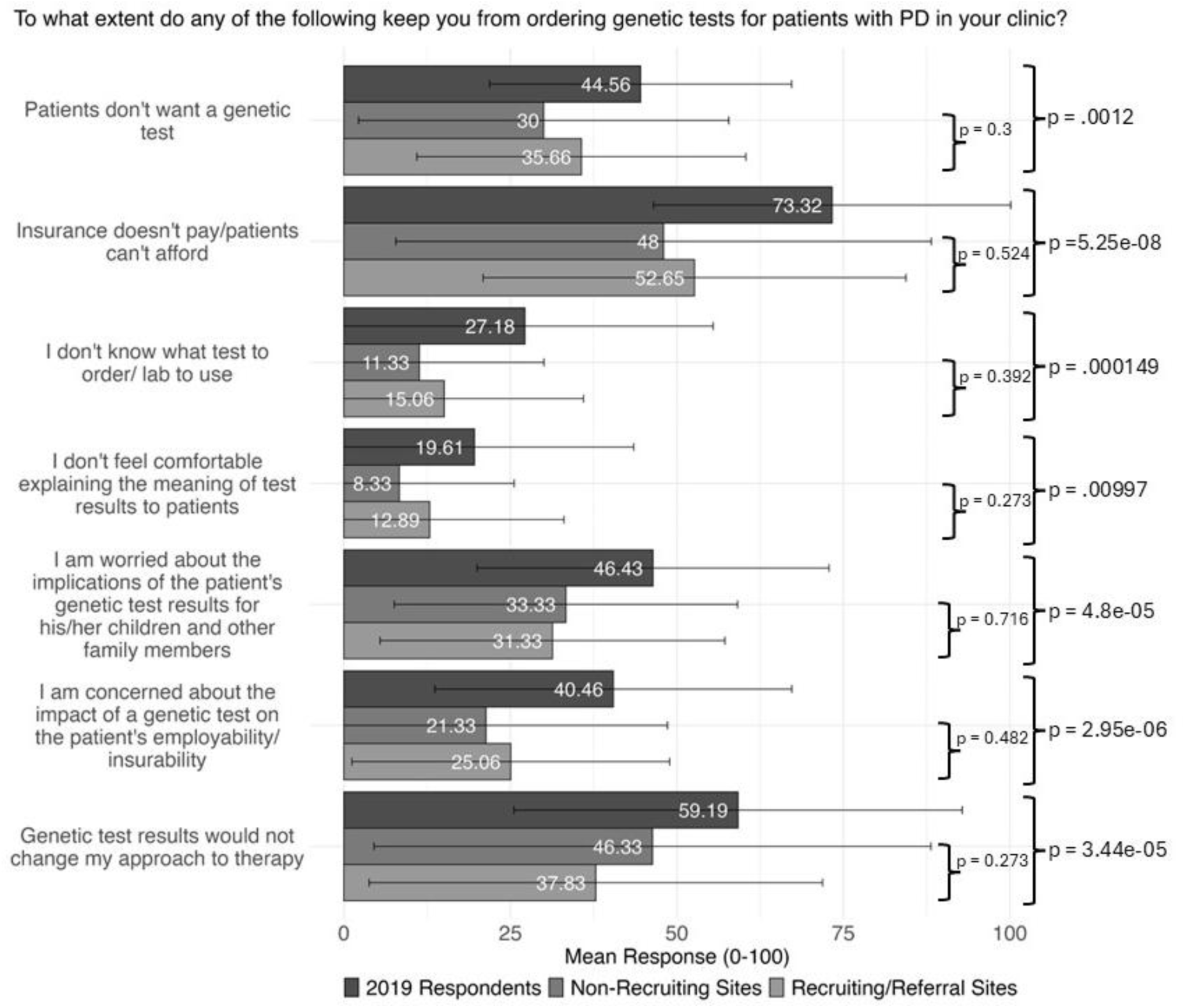
Responses to the question “To what extent do any of the following keep you from ordering genetic tests for patients with Parkinson disease (PD) in your clinic?” across the three groups. Each question was graded on a Likert scale (0 to 100), with each bar representing mean responses and black bars representing standard deviation. All respondents answered each question separately. 2024 respondents are again divided by sites not affiliated with PD GENEration (Non-Recruiting Sites) and sites that are PD GENEration affiliated (Recruiting/Referral Sites). P-value for statistical significance across the three groups determined by ANOVA.

Respondents reporting PD GENEration affiliation (69.7%) were then asked to respond to new questions reflecting on their experiences with PD GENEration (**Figure 3**). Responses to PD GENEration were largely positive, demonstrating participation in PD GENEration has overall led to sites expressing increased comfort with genetic counseling, ease of obtaining genetic test results, improving access to genetic testing and increasing the frequency that genetic tests are ordered. Forty-nine percent of respondents selected 100% when asked how much PD GENEration has improved access to genetic testing for their patients.

**Figure 3:**
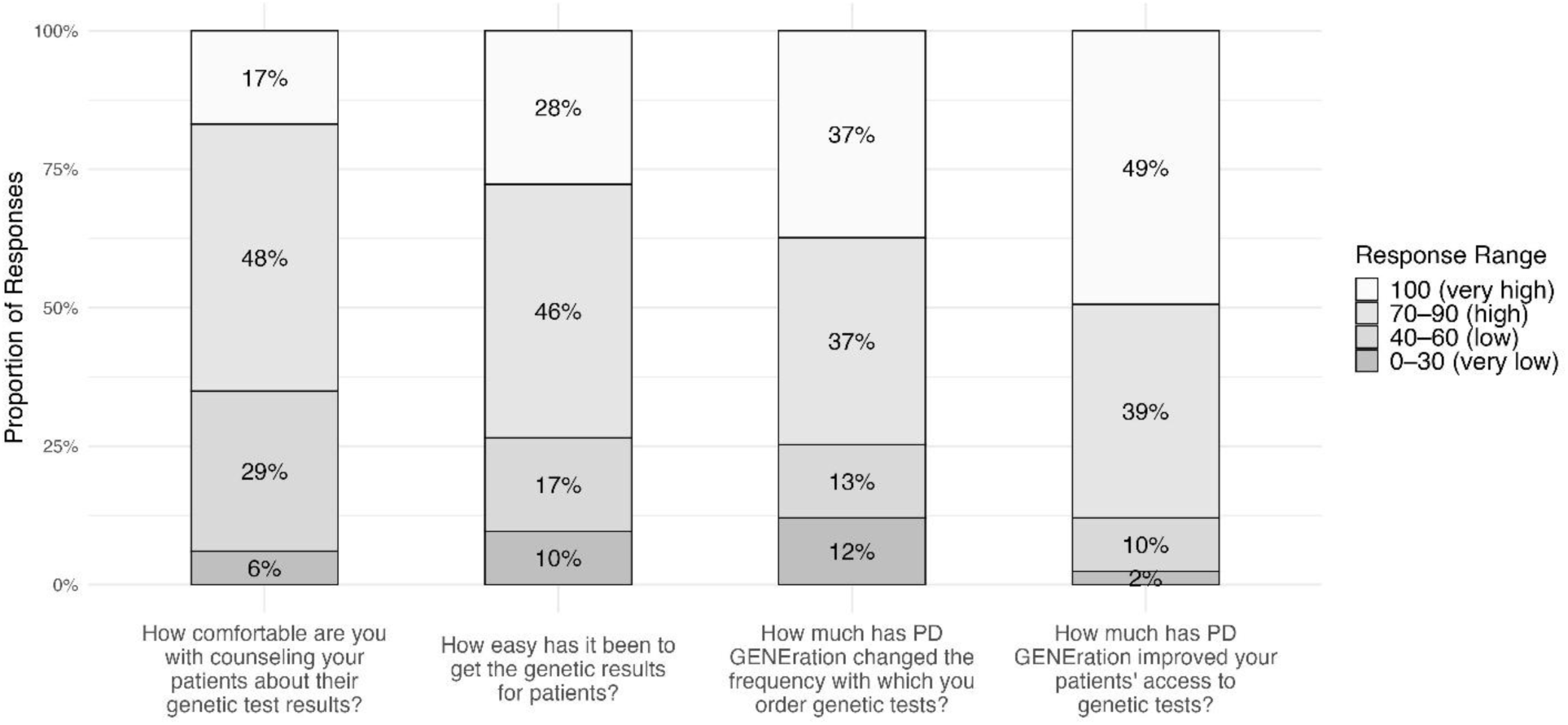
Impact of PD GENEration on genetic testing practices. Survey responses from 83 PD GENEration recruiting and referral sites evaluating the impact of PD GENEration involvement on the following genetic testing practices: comfort with counseling, ease of obtaining results, frequency of test ordering, and improved access to testing. Respondents rated responses on a Likert scale from 0 to 100 and were grouped into very low (0-30%), low (40-60%), high (70-90%) and very high (100%).

### Knowledge and perceived knowledge

To evaluate whether the introduction of PD GENEration positively affected knowledge of PD genetics, clinicians were presented with the same two case scenarios and seven competency questions used in the 2019 survey. Responses to the case scenarios from the 2019 survey, 2024 recruiting/referral PD GENEration sites, and 2024 non-affiliated sites are presented in **Table 1** and **Supplementary Table 2**. As in 2019, 2024 respondents scored well on the basic knowledge questions and reported only a slight and non-significant increase in confidence about their knowledge of genetics.

**Table 1.** Case scenario 1: “Your 65-year-old male Parkinson disease (PD) patient of non-Ashkenazi heritage came to your clinic today to discuss the results of a genetic test that looked for the five most common Gaucher disease– associated GBA1 variants; the test showed that he has one gene with a GBA1 L444P variant and one that is normal for the five variants assayed.” The number and percent of correct, incorrect, and uncertain responses are shown from both the 2019 and 2024 surveys. P values were calculated based on the difference across all groups.

| Case Scenario 1 Responses by Group |  |  |  |  |  |
| --- | --- | --- | --- | --- | --- |
| Question | Group | Correct | Incorrect | Uncertain | $\chi^2$ (p-value) |
| This patient has GBA-associated PD (True statement) | 2019 Respondents | 76<br>(52.4%) | 19<br>(13.1%) | 50 (34.5%) | 12.678 (p = 0.01296) |
|  | 2024 Non-Recruiting sites | 19<br>(63.3%) | 3 (10.0%) | 8 (26.7%) |  |
|  | 2024 Recruiting/Referral | 63<br>(75.9%) | 4 (4.8%) | 16 (19.3%) |  |
| This patient does not have GBA-associated PD, because one copy of the gene is normal (False statement) | 2019 Respondents | 109<br>(75.2%) | 8 (5.5%) | 28 (19.3%) | 6.8857 (p = 0.1421) |
|  | 2024 Non-Recruiting sites | 26<br>(86.7%) | 1 (3.3%) | 3 (10.0%) |  |
|  | 2024 Recruiting/Referral | 73<br>(88.0%) | 1 (1.2%) | 9 (10.8%) |  |
| Deep brain stimulation would NOT be expected to help this patient's motor symptoms (False statement) | 2019 Respondents | 107<br>(73.8%) | 4 (2.8%) | 34 (23.4%) | 8.2097 (p = 0.08419) |
|  | 2024 Non-Recruiting sites | 23<br>(76.7%) | 3 (10.0%) | 4 (13.3%) |  |
|  | 2024 Recruiting/Referral | 63<br>(75.9%) | 8 (9.6%) | 12 (14.5%) |  |
| Each of this patient's biological children has a 50% chance of developing Parkinson disease (False statement) | 2019 Respondents | 119<br>(82.1%) | 9 (6.2%) | 17 (11.7%) | 6.7654 (p = 0.1488) |
|  | 2024 Non-Recruiting sites | 19<br>(67.9%) | 6 (21.4%) | 3 (10.7%) |  |
|  | 2024 Recruiting/Referral | 64<br>(77.1%) | 9 (10.8%) | 10 (12.0%) |  |
| Each of this patient's biological children has a 50% chance of developing Gaucher disease (False statement) | 2019 Respondents | 116<br>(80.0%) | 3 (2.1%) | 26 (17.9%) | 2.6782 (p = 0.613) |
|  | 2024 Non-Recruiting sites | 24<br>(80.0%) | 2 (6.7%) | 4 (13.3%) |  |
|  | 2024 Recruiting/Referral | 68<br>(82.9%) | 3 (3.7%) | 11 (13.4%, 1 NA) |  |

Case scenario 1 asked about a patient with a single *GBA1 L444P* variant. When asked whether the participant had *GBA1*-associated PD (correct answer), only 52.4% of participants in 2019 replied in the affirmative, compared to 76.8% of participants in 2024 in PD GENEration affiliated sites and 63.3% of participants in non-affiliated sites (overall χ² p value = 0.013). None of the other knowledge questions differed significantly between 2019 and 2024 (Table 1); however, the percent of correct responses in the 2019 survey was already high.

Case scenario 2 asked about a participant with the *G2019S LRRK2* genetic variant. More respondents in 2024 answered the following false statement correctly “this patient is more likely to have cognitive impairment sooner than a patient with idiopathic PD” (Supplementary Table 2, p = 0.0006) than in 2019. With subsequent questions, there was no change in the already high accuracy of responses from 2019.

Overall, on a Likert scale of 0-100, with 100 being very confident, there was no significant difference in survey participants’ confidence “in their knowledge of the impact of genetic variants on PD phenotypic manifestations,” (mean 47.6, SD = 26.3 in 2019, vs 50.47, SD = 25.16 in 2024). Similarly, feeling “up to date on genetically targeted experimental therapeutics on *GBA1* and *LRRK2-*related trials,” was unchanged between 2019 (mean 48.8, SD = 28.3) and 2024 (mean 48.98, SD = 24.49). Nor was there a difference in feeling “comfortable counseling their PD patients on *GBA1* and *LRRK2* testing,” (mean 51.6, SD = 27.5 in 2019 vs 52.32, SD = 26.37 in 2024).

## DISCUSSION

Compared to 2019, clinicians responding to the 2024 survey reported significantly greater comfort with genetic testing, fewer perceived barriers, and increased testing volumes— particularly at PD GENEration sites. There was a marked reduction in perceived barriers to genetic testing and an improvement in both perceived access to testing and clinicians’ willingness to send genetic tests. We were surprised to find that even within the sites that were not affiliated with PD GENEration, the perceived barriers to genetic testing have reduced, suggesting that the effect of free genetic testing has had a broader impact on the Parkinson’s community.

Limitations of our study include the fact that the survey was sent only to PSG members who are already experts in Parkinson’s disease and therefore is not representative of all clinicians in North America or even all neurologists. However, the survey was designed to obtain and compare similar results between 2019 and 2024 in order to determine the effects of PD GENEration in a controlled manner. Another limitation was the limited information on the effect of ROPAD on PSG sites. As only 11 PSG sites are involved in ROPAD, we did not separate out this group. A third limitation is the fact that a smaller number of participants completed the 2024 survey compared to 2019. It is unclear whether this could have biased our results either positively or negatively.

Although PD GENEration, the PSG, and GP2, have made efforts to educate the community around genetics, genetic testing, and genetic counseling ^6,19,20^, there remains a clear need for continuing education. Our survey results suggest that attitudes towards genetic testing are changing within the Parkinson’s community and improving as a result of increased access to genetic testing through research protocols such as PD GENEration, thereby setting the stage for the advent of precision medicine trials for genetic forms of PD.

## DATA AVAILABILITY

The data and code underlying this study are not publicly available due to privacy concerns as many respondents voluntarily provided contact information. Data and code can be provided upon request. Requests for access should be directed towards the corresponding author.

## Data Availability

The data and code underlying this study are not publicly available due to privacy concerns as many respondents voluntarily provided contact information. Data and code can be provided upon reasonable request. Requests for access should be directed towards the corresponding author.

## ACKNOWLEDGEMENTS

We thank the Parkinson’s Study Group, Sarah Sperling, and Donna Moszkowicz for their help in implementing and distributing the questionnaire.

## FUNDING STATEMENT

No funding was received for this research.

## AUTHOR CONTRIBUTIONS

Conceptualization: R.N.A., J.C.B., M.N., A.M.W.; Data curation: J.W., V.I.; Formal analysis: J.W., V.I; Project administration: M.E.C.; Visualization: J.W., V.I.; Writing-original draft: J.W., A.M.W.; Writing-review & editing: R.N.A., J.A.L., M.N., J.C.B., A.N., K.G.G.

## ETHICS DECLARATION

This protocol was reviewed and approved by the Advarra IRB. Due to the minimal risk involved with this study, participants provided informed consent through acceptance of the study invitation and completing an electronic consent.

## CONFLICT OF INTEREST

A.M.W. has received research funding from NIH/NIA, the Parkinson’s Foundation, participated in clinical trials sponsored by Amylyx Pharmaceuticals, Roche/Genentech, Biogen, Ono pharmaceuticals, Bial pharmaceuticals, research funding from BioSensics, and has received consulting fees from Genentech, Ono Pharmaceuticals, and Arvinas. R.N.A. research is funded by the Michael J. Fox Foundation, the Silverstein Foundation, the Parkinson’s Foundation and the Aufzien Family Center for the Prevention and Treatment of Parkinson’s Disease. He received consultation fees from Bexxion, Biogen, Biohaven, Capsida, Gain Therapeutics, Genzyme/Sanofi, Janssen, SK Biopharmaceuticals, Takeda and Vanqua Bio. M.N. has received grant support for Centers of Excellence from the Parkinson Foundation, and compensation for PDGENE Steering Committee, ACTIVATE trial sponsored by Bial, and KINECT-HD trial sponsored by Neurocrine. M.N. is Co-chair, Genetics and Environment Working group and Member, mentorship committee, Parkinson Study Group. The other authors declare no conflicts of interest.

## Footnotes

* The Sanofi MOVES-PD trial^13^ (NCT02906020) was the sole precision medicine clinical trial active in the US in 2019. Active trials for PD in 2024 include BIAL ACTIVATE (NCT05819359)^14^, Prevail PROPEL (NCT04127578)^15^, Denali BEACON (NCT06602193)^16^, Neuron 23 NEULARK (NCT06680830)^17^, and Biogen REASON (NCT03976349)^18^.

## SUPPLEMENTARY MATERIAL

**Table 2.** Case scenario 2 stated: “A 60-year-old male Parkinson disease (PD) patient of Ashkenazi background with a positive family history of Parkinson disease comes to your clinic to discuss the results of their LRRK2 genetic test, which shows a G2019S variant on one copy of the gene, with the other copy showing no variants”. The number and percent of correct, incorrect, and uncertain responses are shown from both the 2019 and 2024 surveys. P-values were calculated based on the difference across all groups.

| Case Scenario 2 Responses by Group |  |  |  |  |  |
| --- | --- | --- | --- | --- | --- |
| Question | Group | Correct | Incorrect | Uncertain | $\chi^2$ (p-value) |
| This patient has LRRK2-associated PD (True statement) | 2019 Respondents | 123<br>(84.8%) | 3 (2.1%) | 19<br>(13.1%) | 9.9508 (p = 0.04126) |
|  | 2024 Non-Recruiting sites | 23<br>(76.7%) | 4 (13.3%) | 3 (10%) |  |
|  | 2024 Recruiting/Referral | 72<br>(85.7%) | 2 (2.4%) | 10<br>(11.9%) |  |
| This patient does not have LRRK2-associated PD, because one copy of the gene is normal (False statement) | 2019 Respondents | 129<br>(89.0%) | 5 (3.4%) | 11 (7.6%) | 0.56519 (p = 0.9669) |
|  | 2024 Non-Recruiting sites | 27 (90%) | 1 (3.3%) | 2 (6.7%) |  |
|  | 2024 Recruiting/Referral | 73<br>(88.0%) | 2 (2.4%) | 8 (9.6%) |  |
| This patient is more likely to have cognitive impairment sooner than a patient with idiopathic PD (False statement) | 2019 Respondents | 91<br>(62.8%) | 20<br>(13.8%) | 34<br>(23.4%) | 14.443 (p = 0.006006) |
|  | 2024 Non-Recruiting sites | 26<br>(86.7%) | 2 (6.7%) | 2 (6.7%) |  |
|  | 2024 Recruiting/Referral | 68<br>(81.9%) | 8 (9.6%) | 7 (8.4%) |  |
| Deep brain stimulation would NOT be expected to help this patient's motor symptoms (False statement) | 2019 Respondents | 122<br>(84.1%) | 2 (1.4%) | 21<br>(14.5%) | 7.6112 (p = 0.1069) |
|  | 2024 Non-Recruiting sites | 28<br>(93.3%) | 1 (3.3%) | 1 (3.3%) |  |
|  | 2024 Recruiting/Referral | 77<br>(92.8%) | 2 (2.4%) | 4 (4.8%) |  |
| Each of this patient's biological children has a 50% chance of developing Parkinson disease (False statement) | 2019 Respondents | 108<br>(74.5%) | 20<br>(13.8%) | 17<br>(11.7%) | 12.959 (p = 0.01148) |
|  | 2024 Non-Recruiting sites | 19<br>(63.3%) | 11<br>(36.7%) | 0 (0%) |  |
|  | 2024 Recruiting/Referral | 64<br>(77.1%) | 14<br>(16.9%) | 5 (6.0%) |  |
| Each of this patient's biological children has a 50% chance of developing Gaucher disease (False statement) | 2019 Respondents | 133<br>(91.7%) | 3 (2.1%) | 9 (6.2%) | 10.731 (p = 0.02976) |
|  | 2024 Non-Recruiting sites | 29<br>(96.7%) | 1 (3.3%) | 0 (0%) |  |
|  | 2024 Recruiting/Referral | 77<br>(92.8%) | 6 (7.2%) | 0 (0%) |  |
| Testing for the G2019S variant is available through direct-to-consumer testing (True statement) | 2019 Respondents | 89<br>(61.4%) | 1 (0.7%) | 55<br>(37.9%) | 2.5369 (p = 0.638) |
|  | 2024 Non-Recruiting sites | 15 (50%) | 1 (3.3%) | 14<br>(46.7%) |  |
|  | 2024 Recruiting/Referral | 50<br>(60.2%) | 1 (1.2%) | 32<br>(38.6%) |  |

---

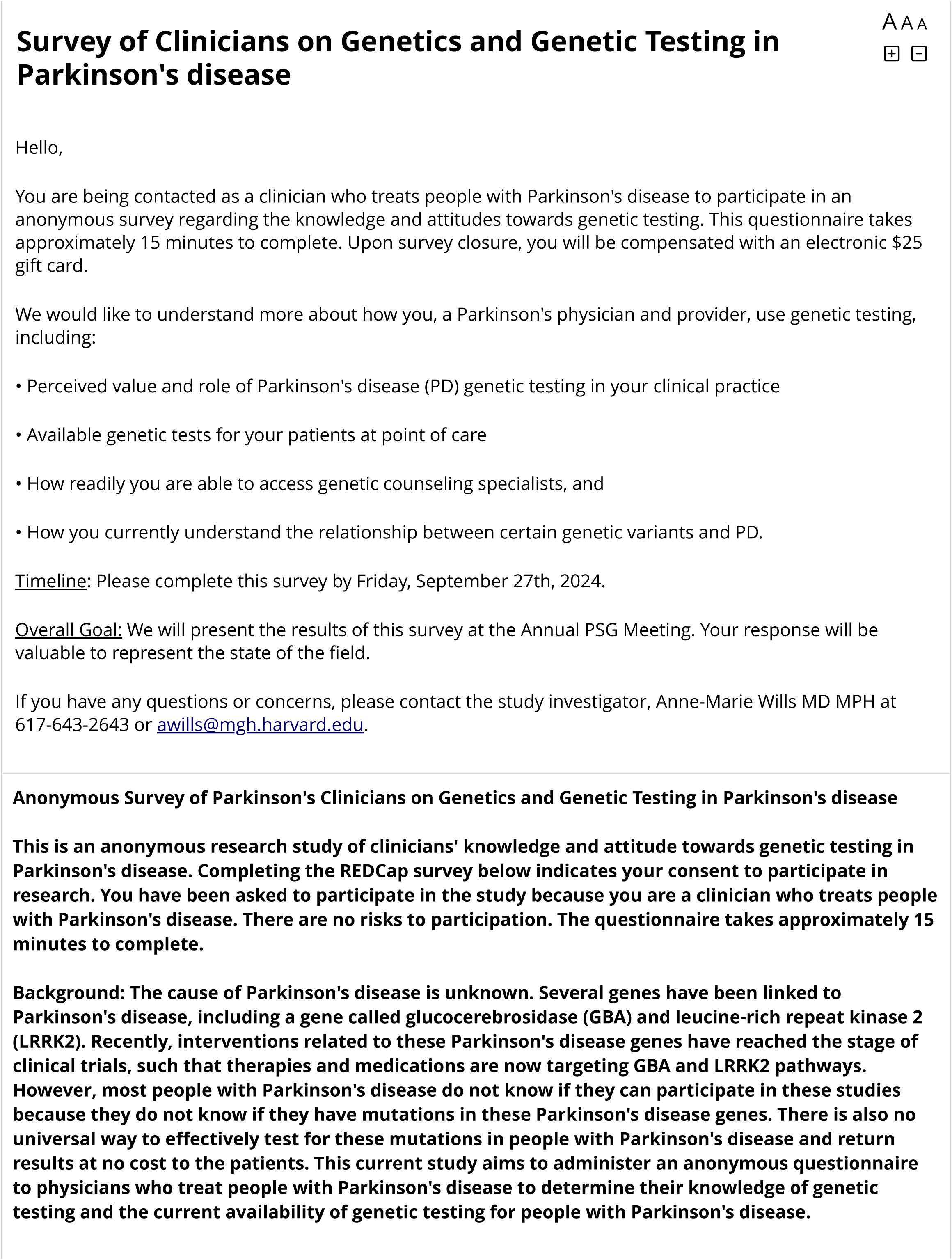
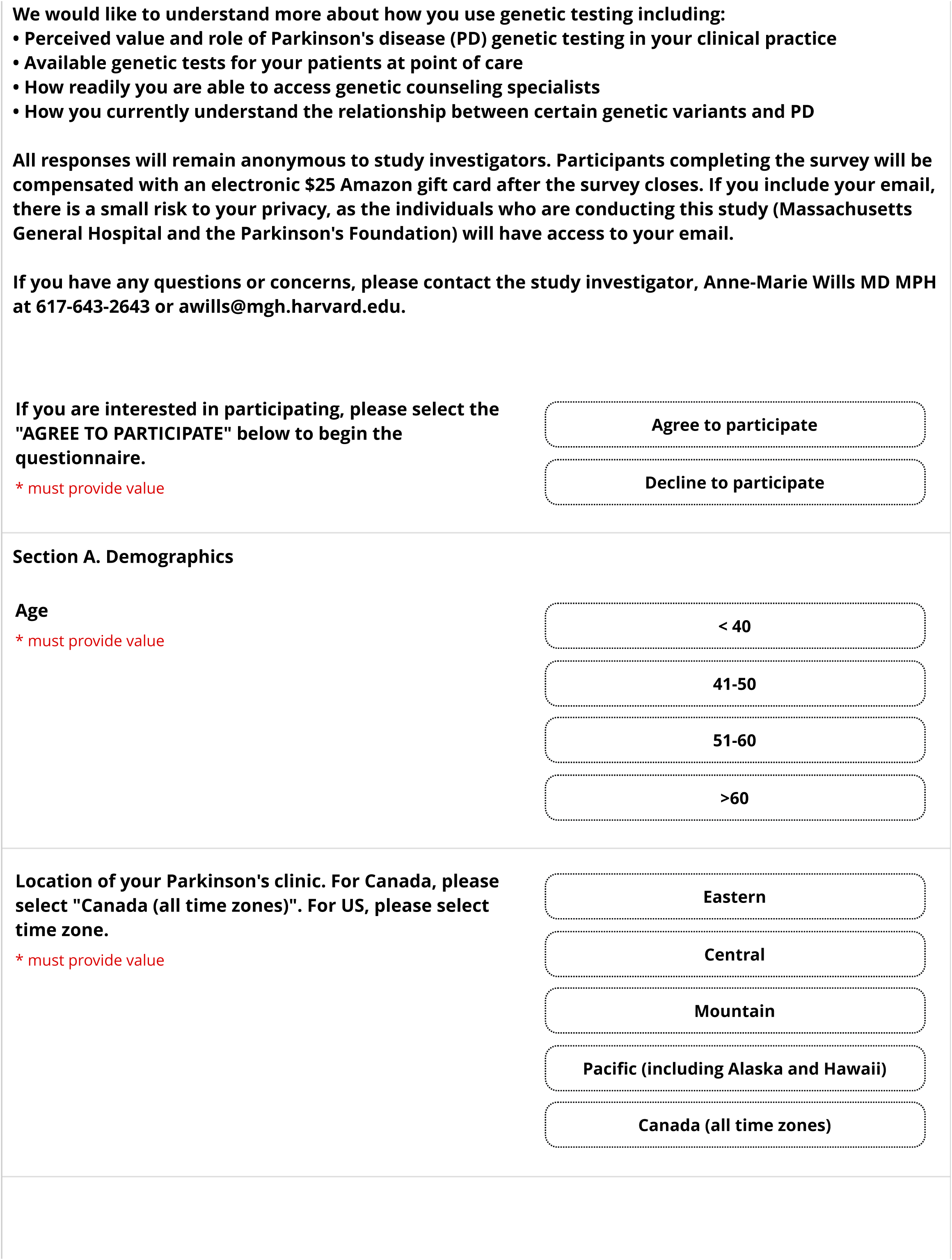
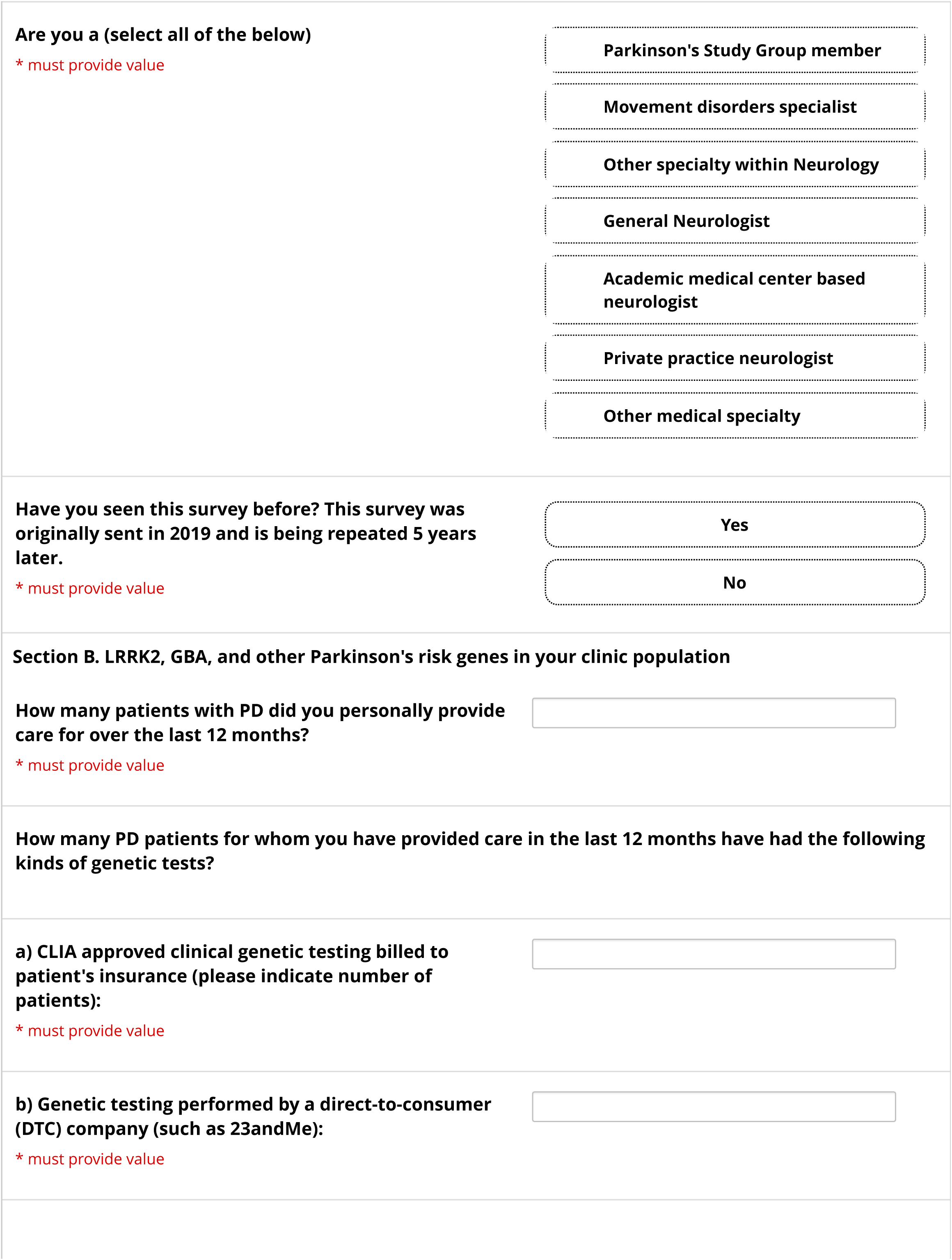
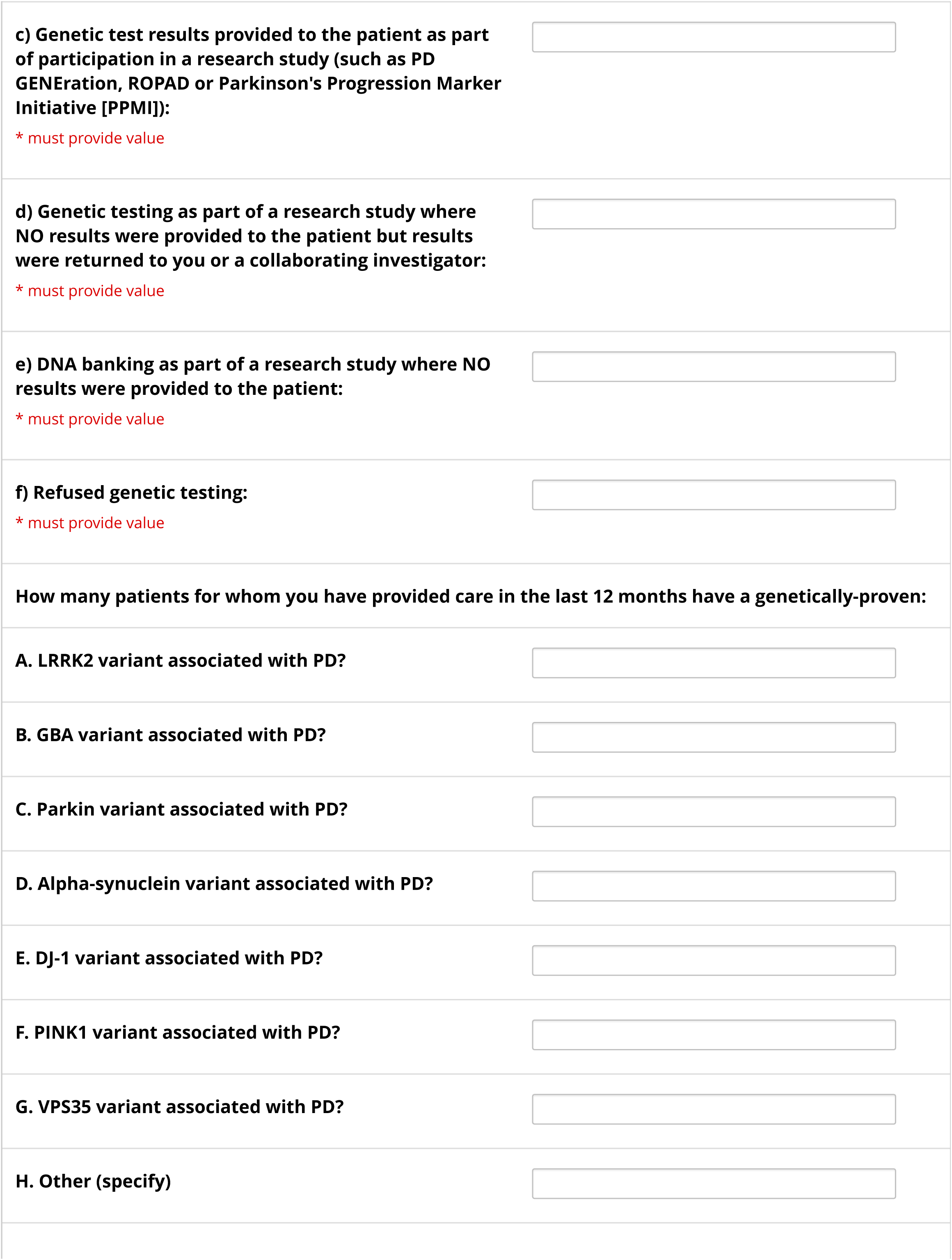
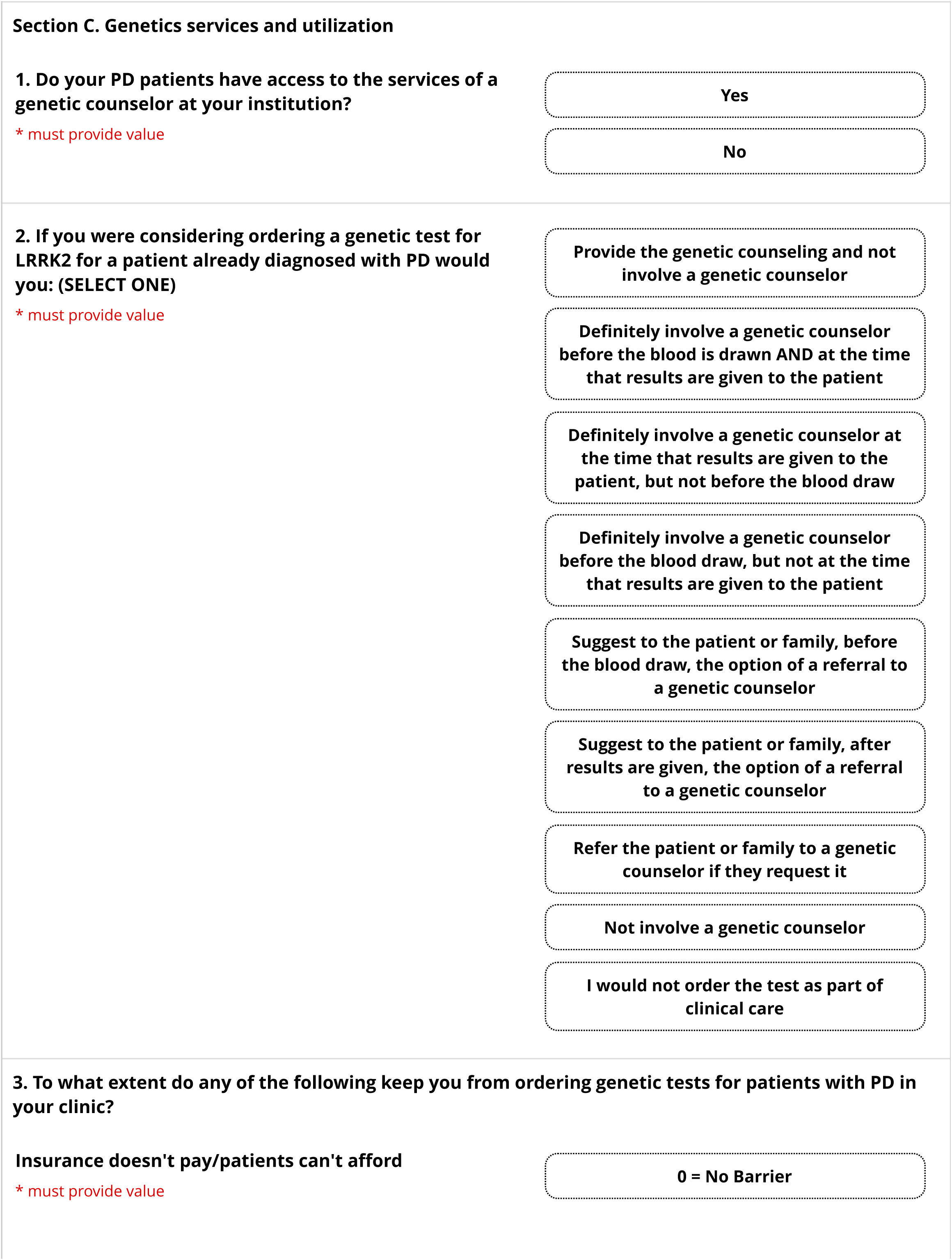
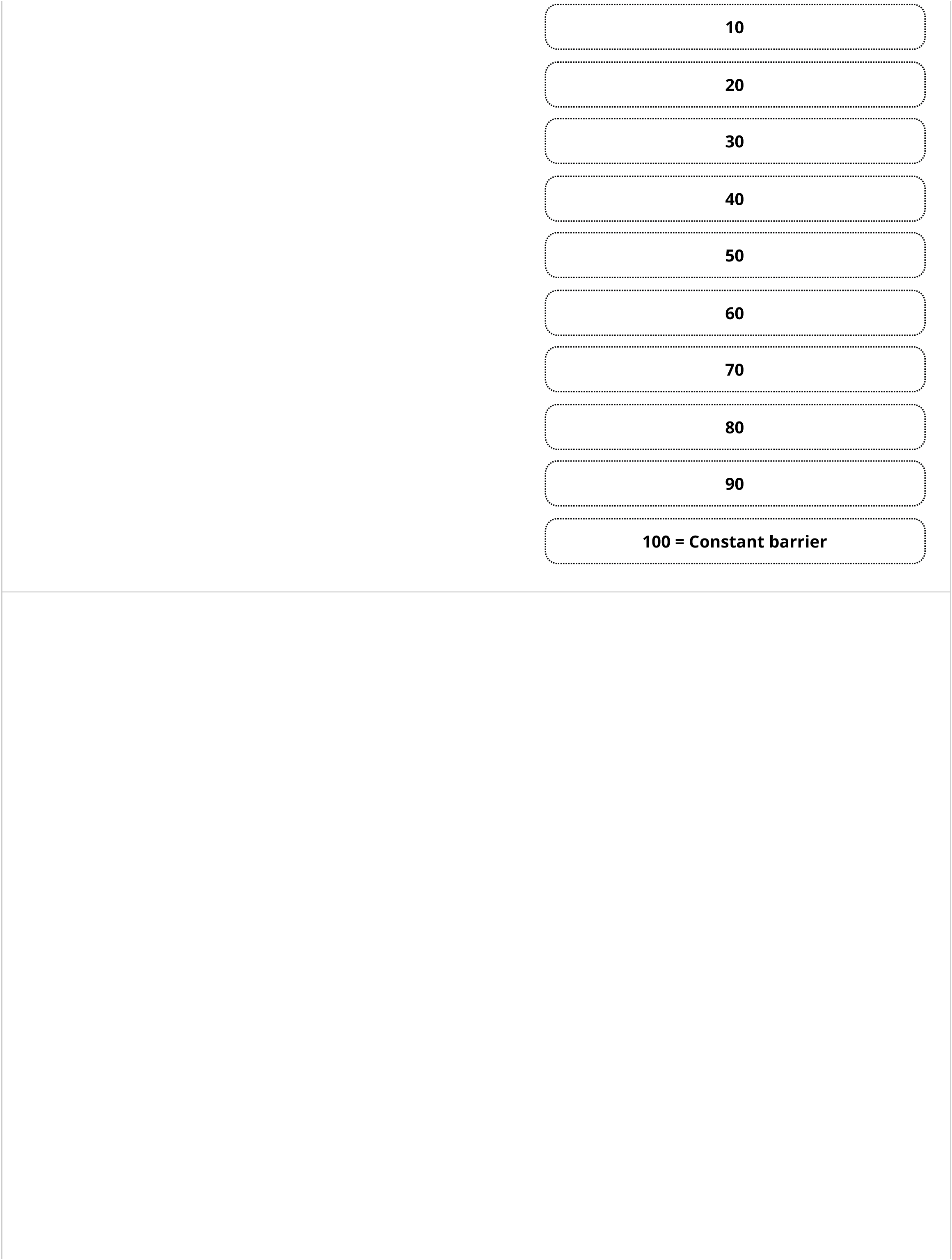
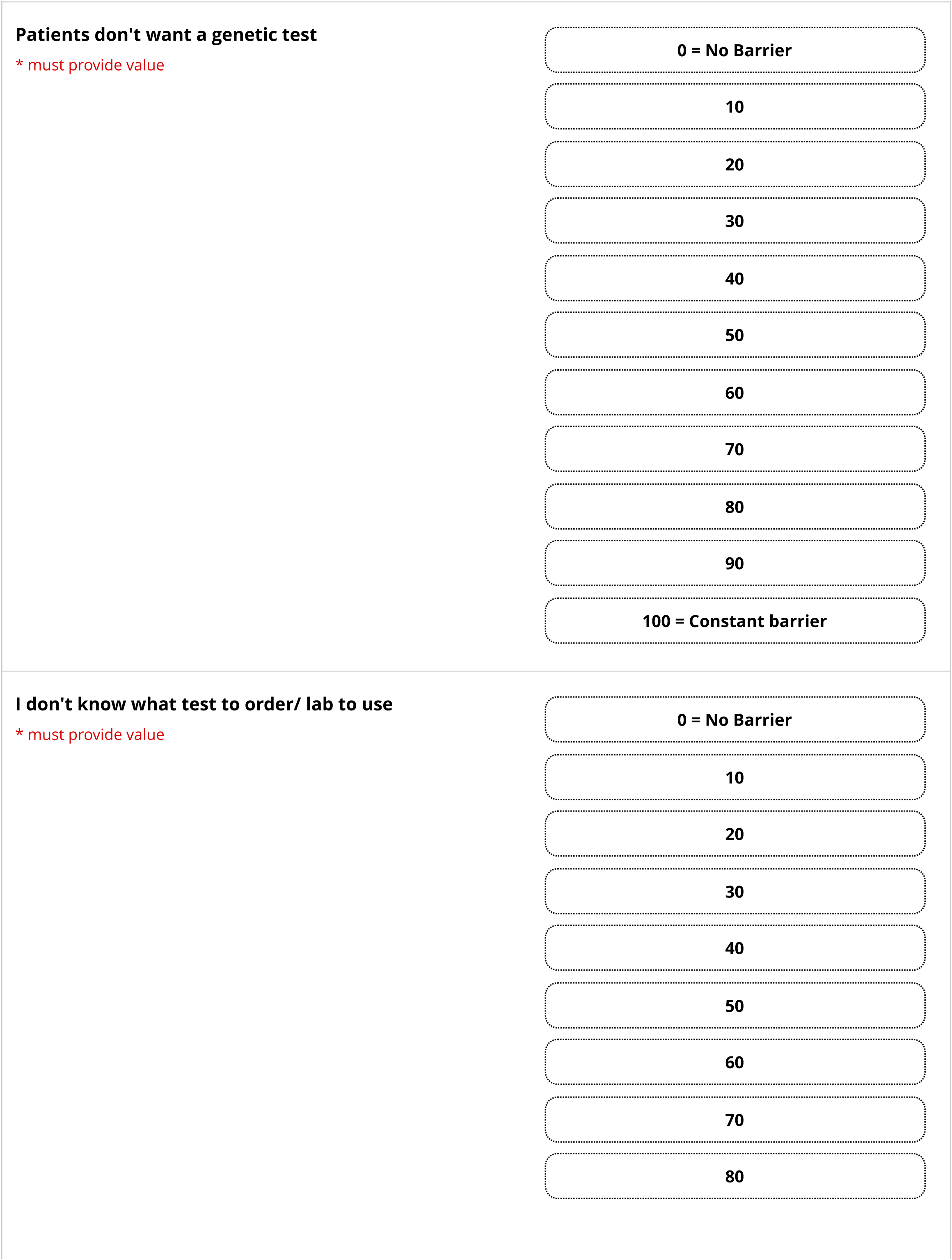
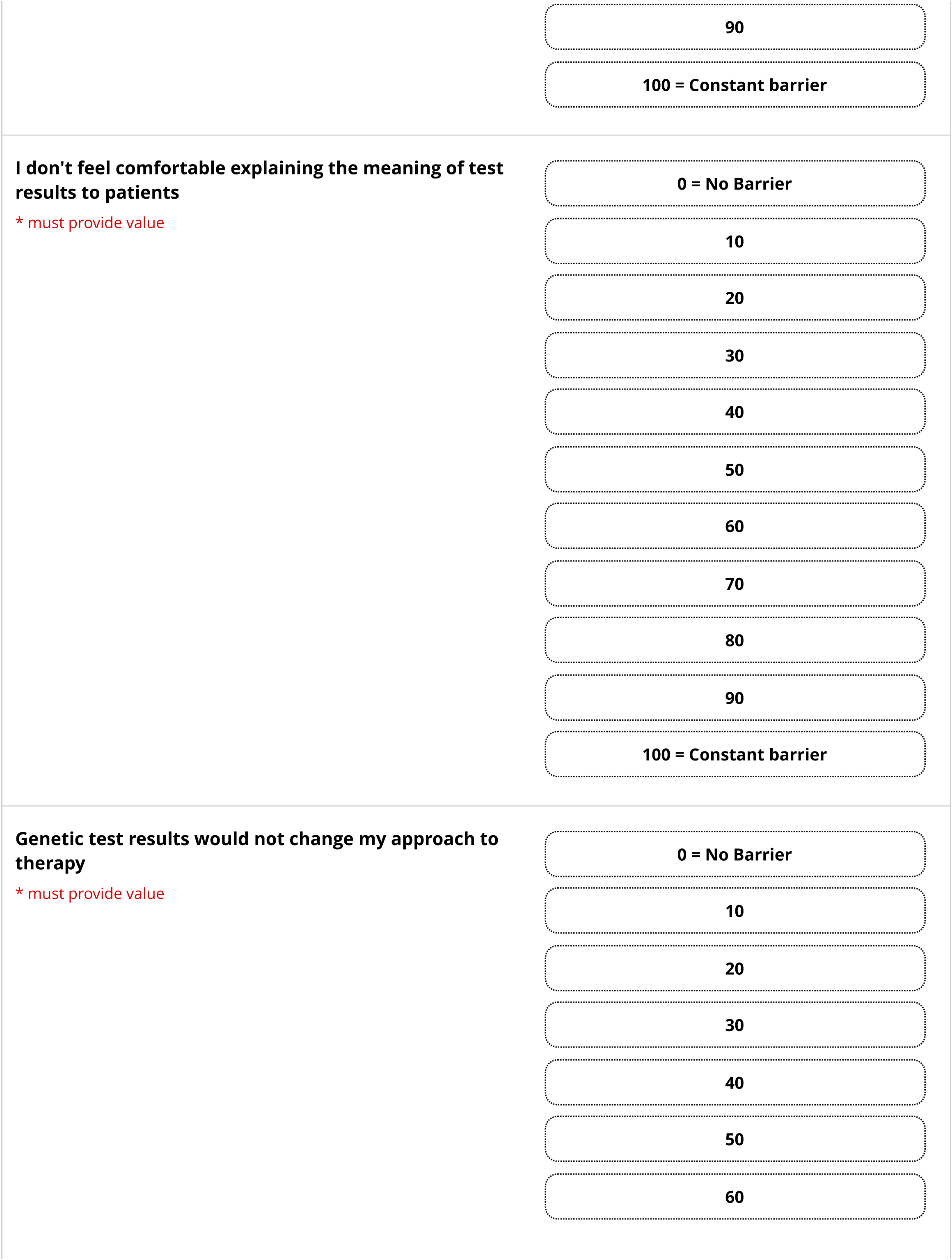
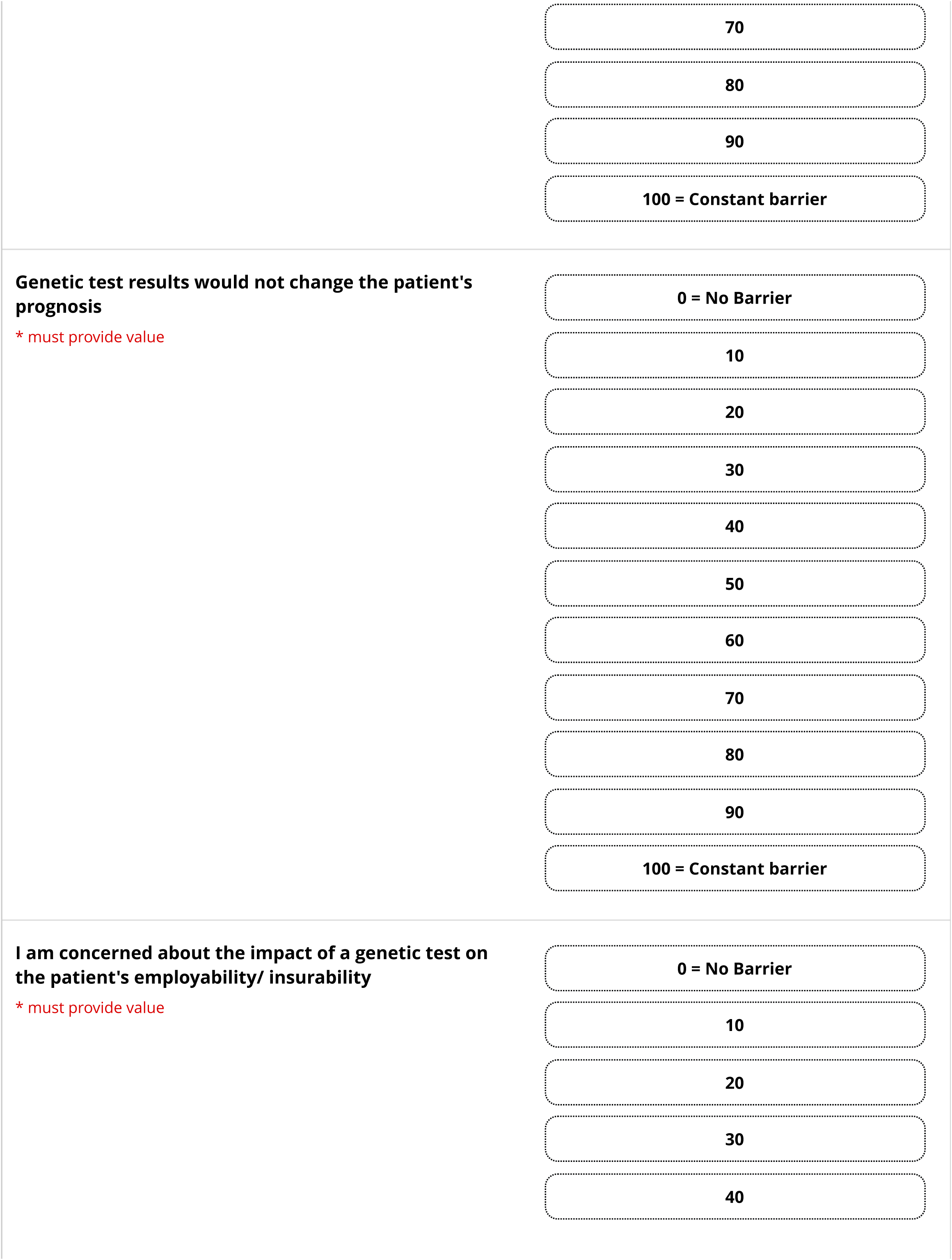
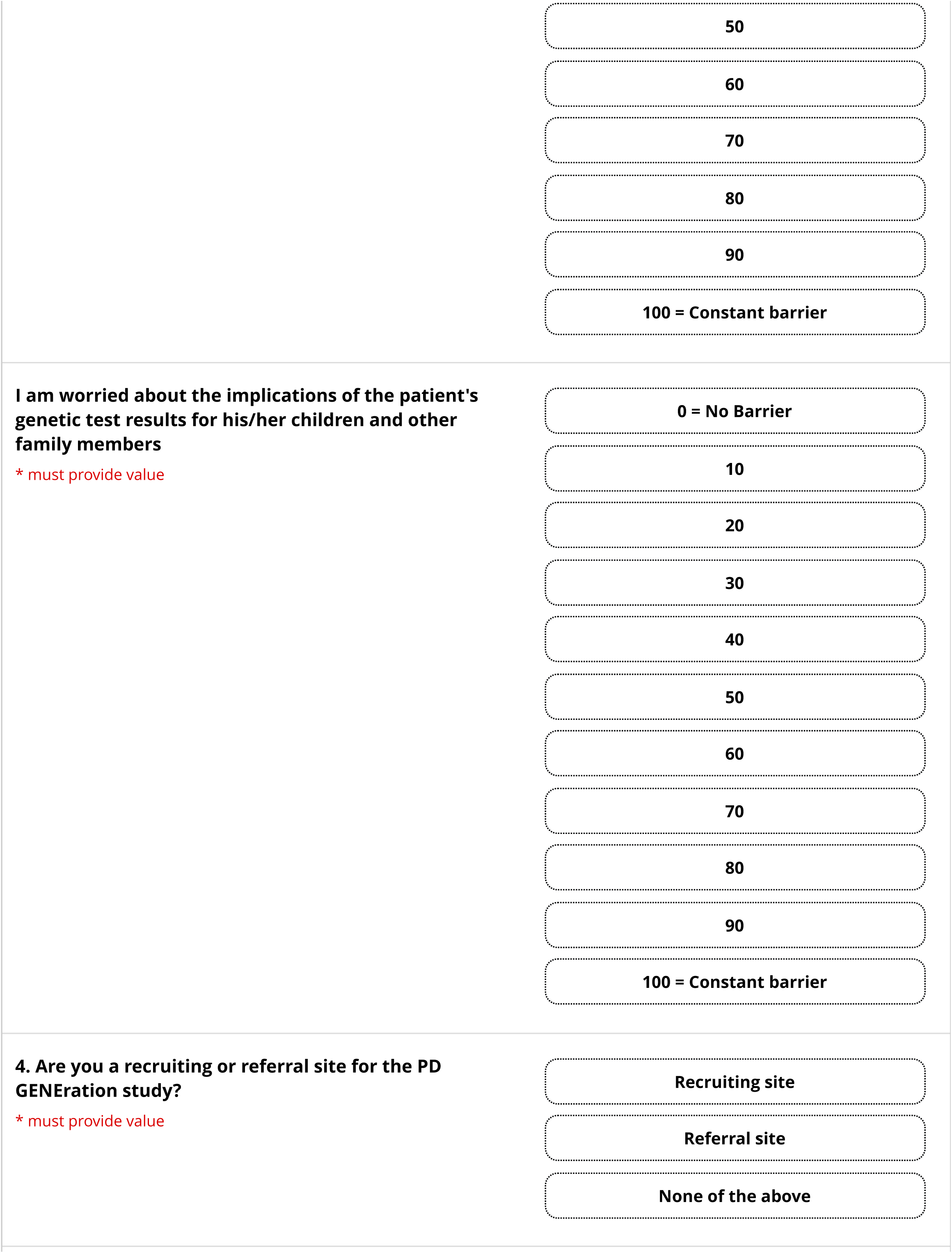
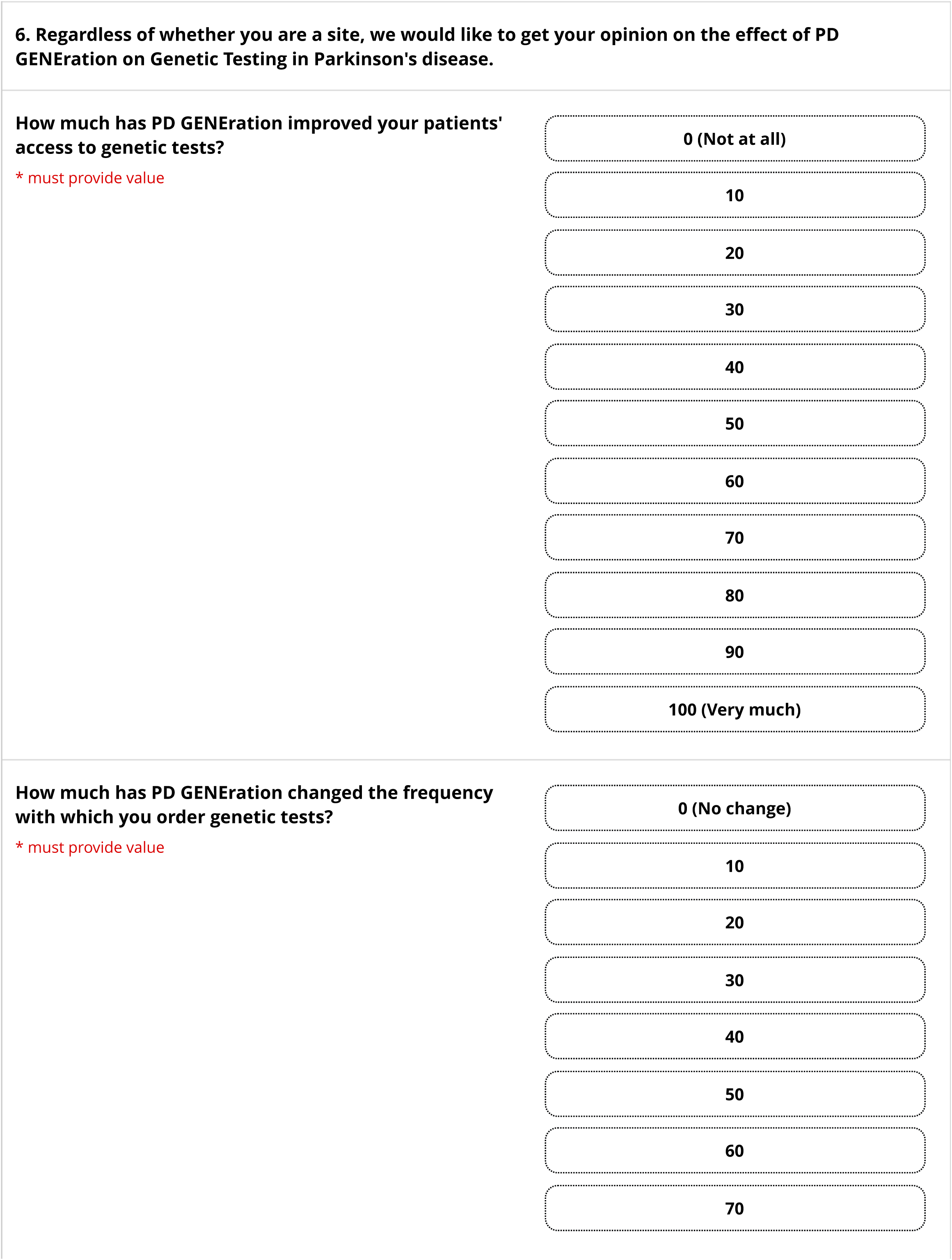
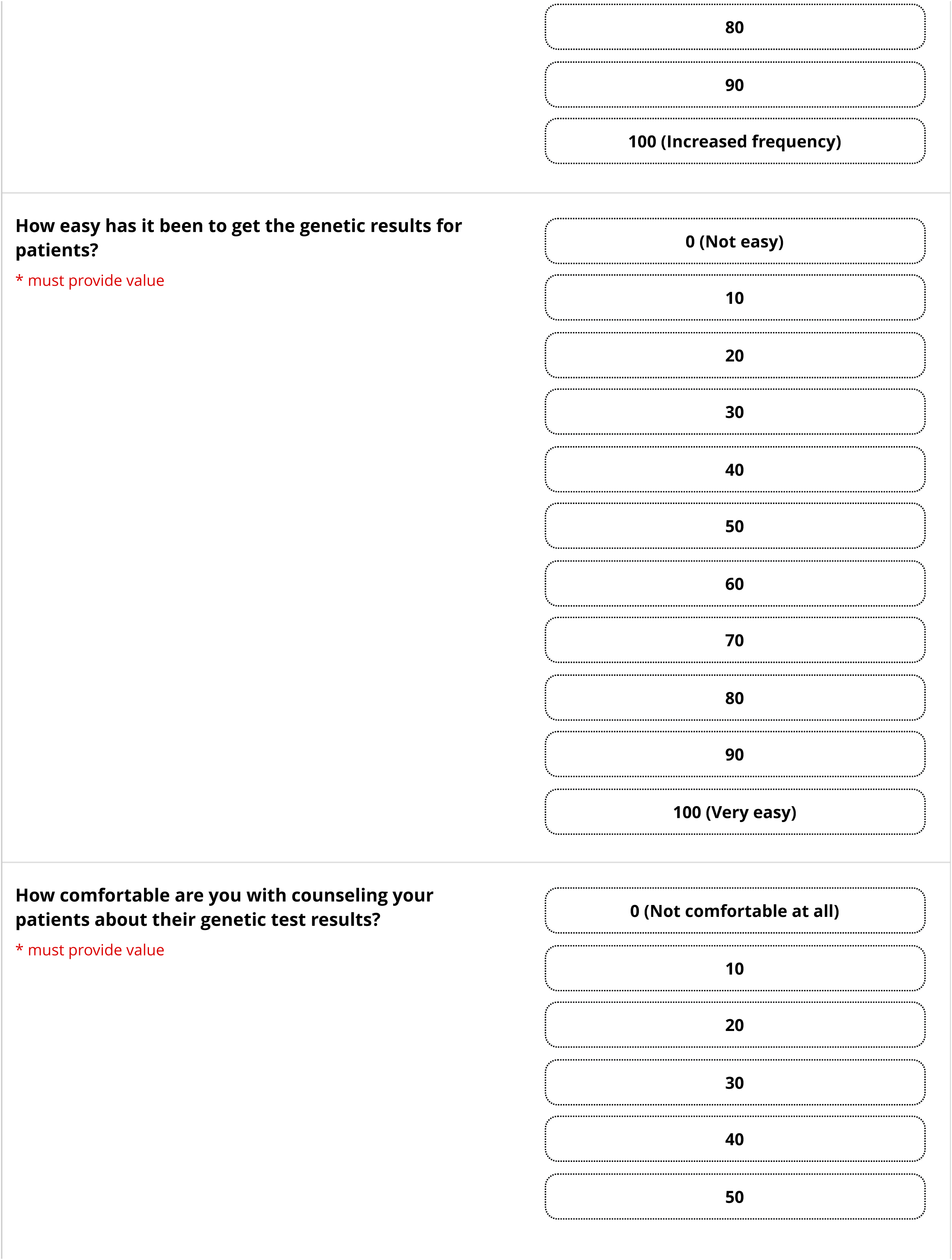
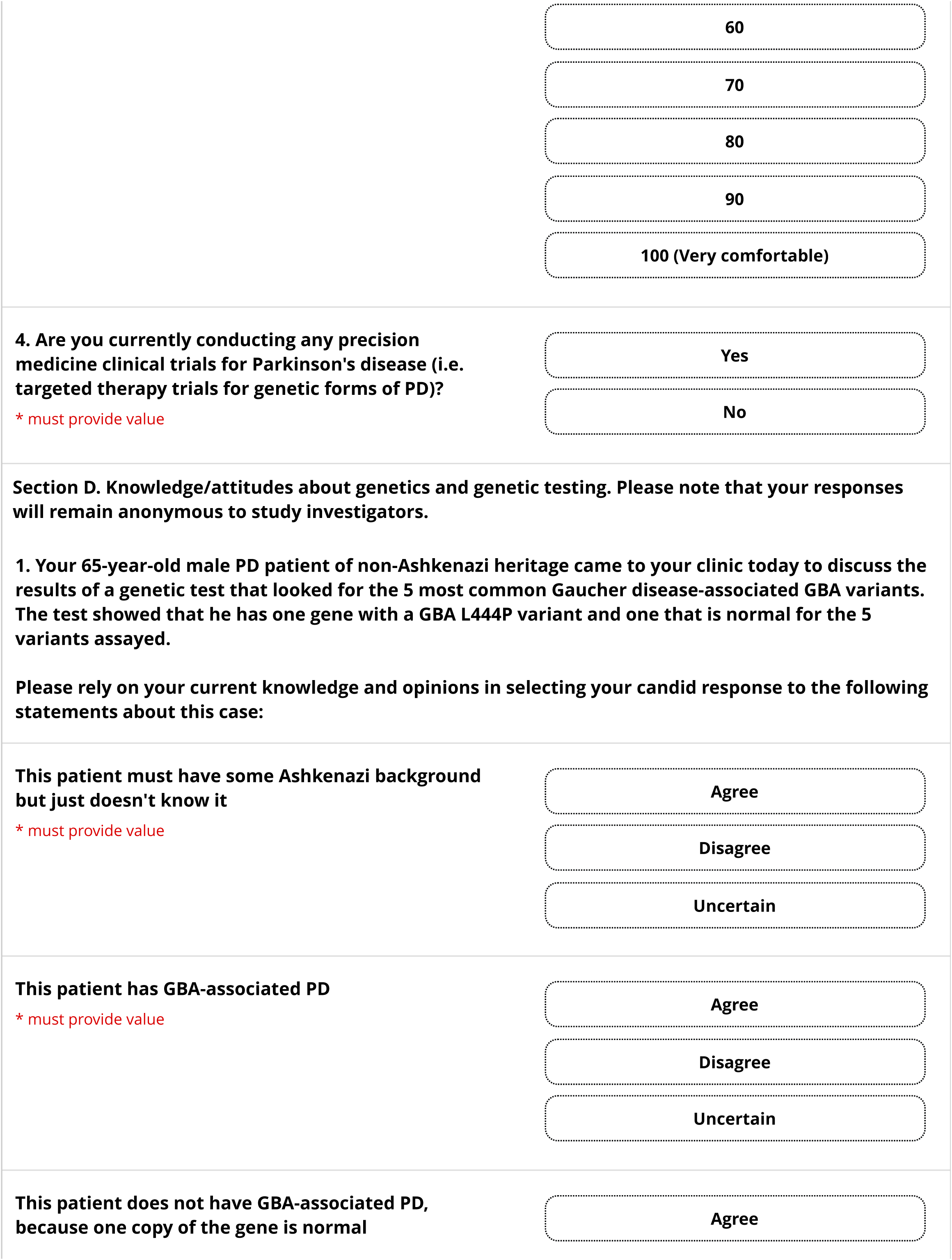
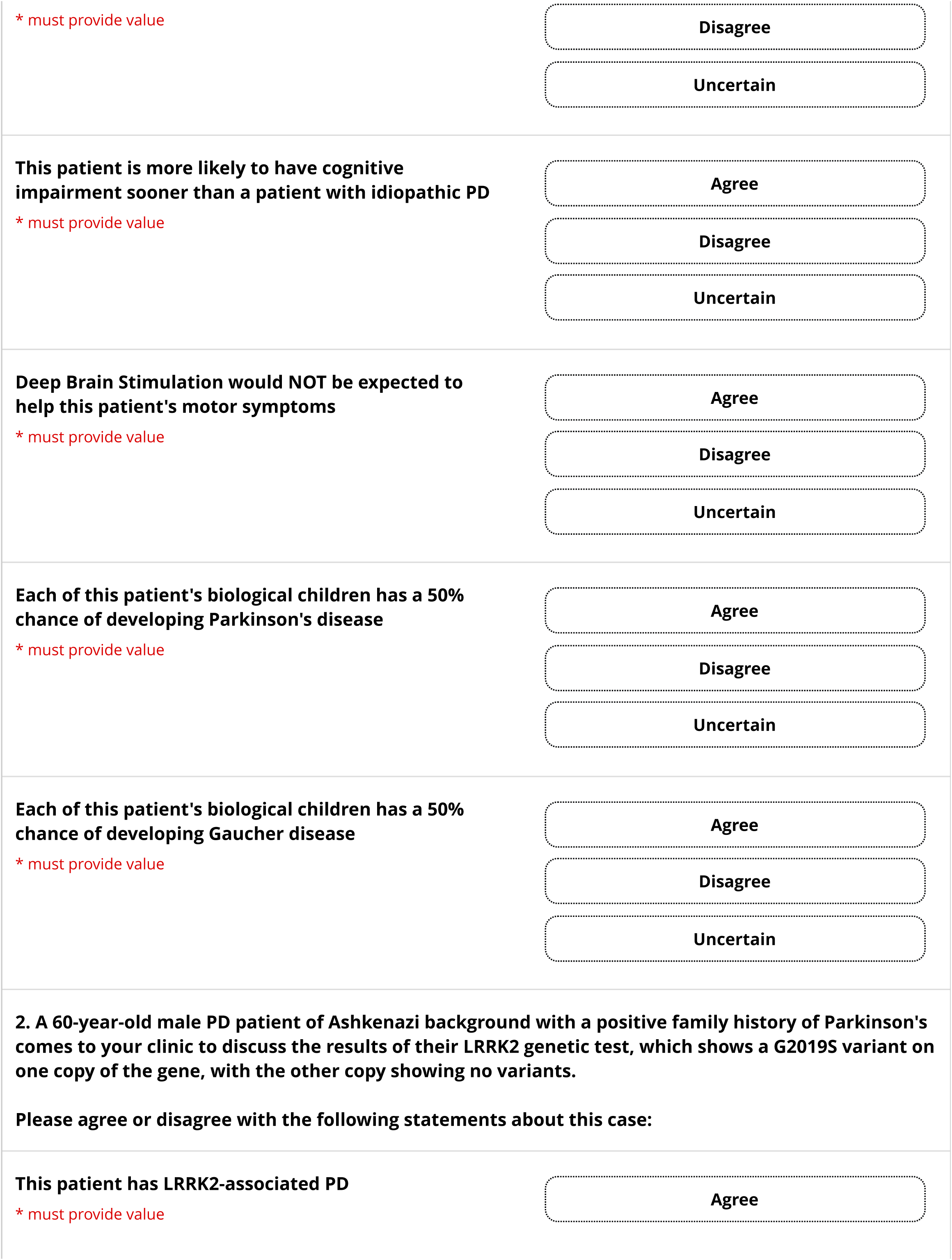
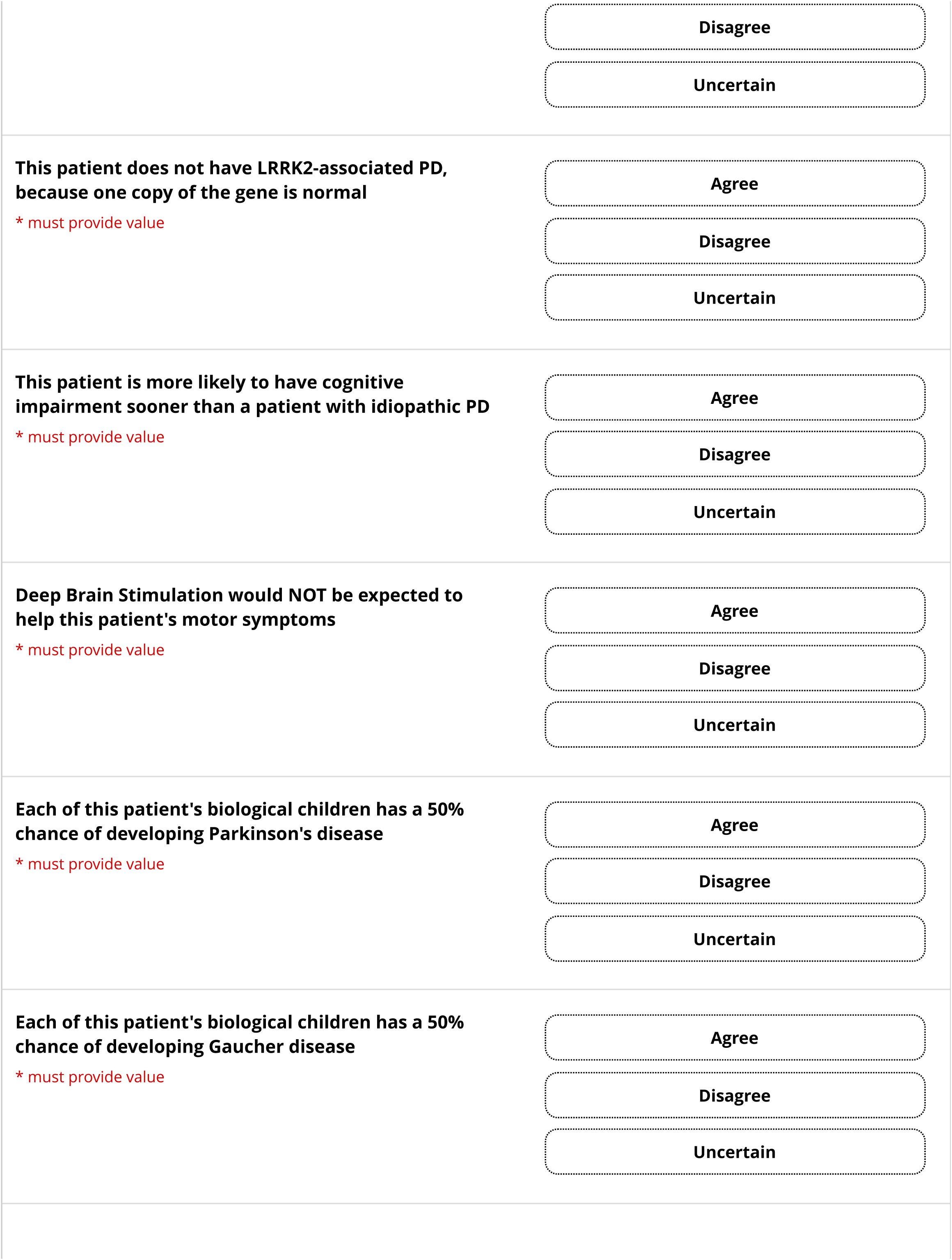
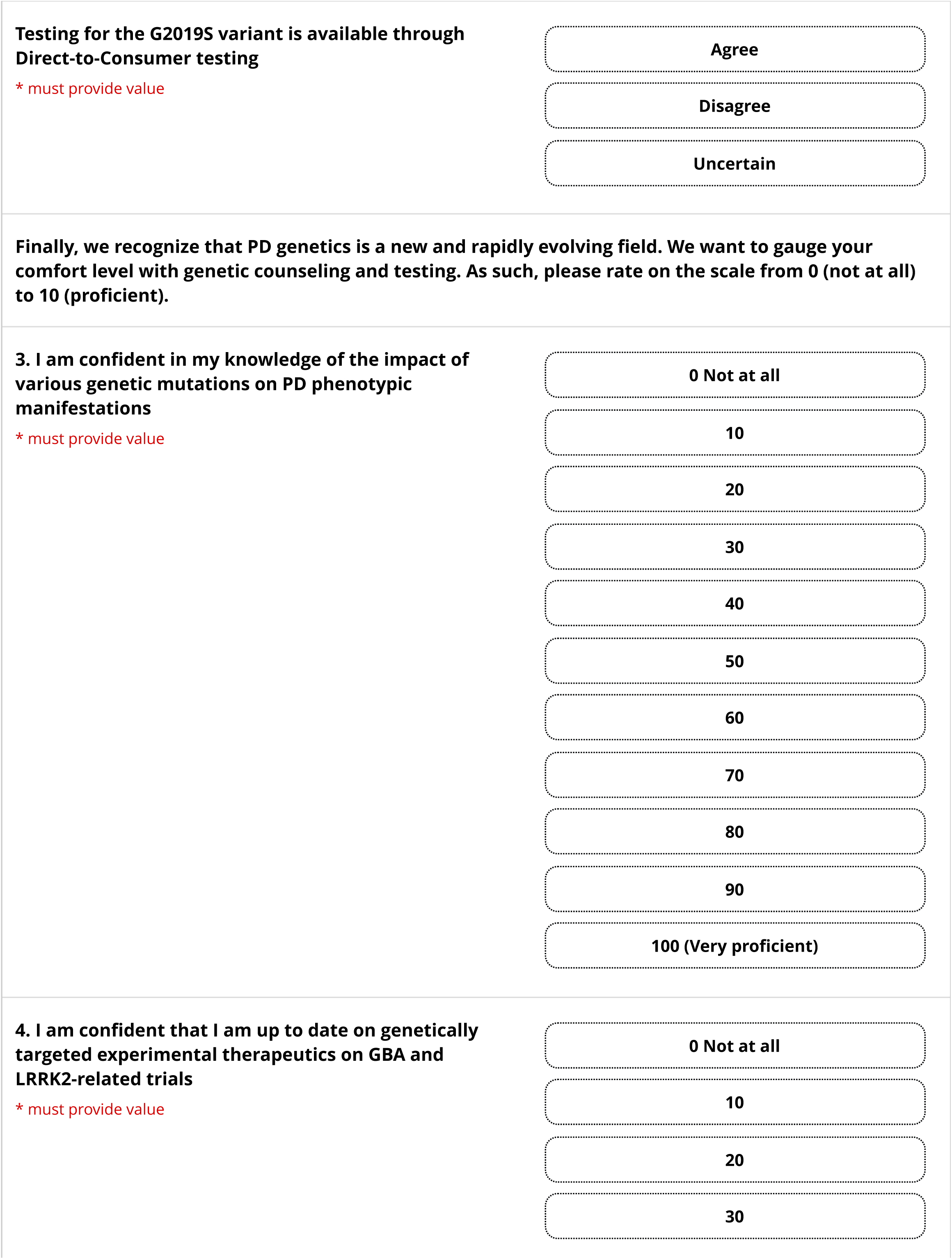
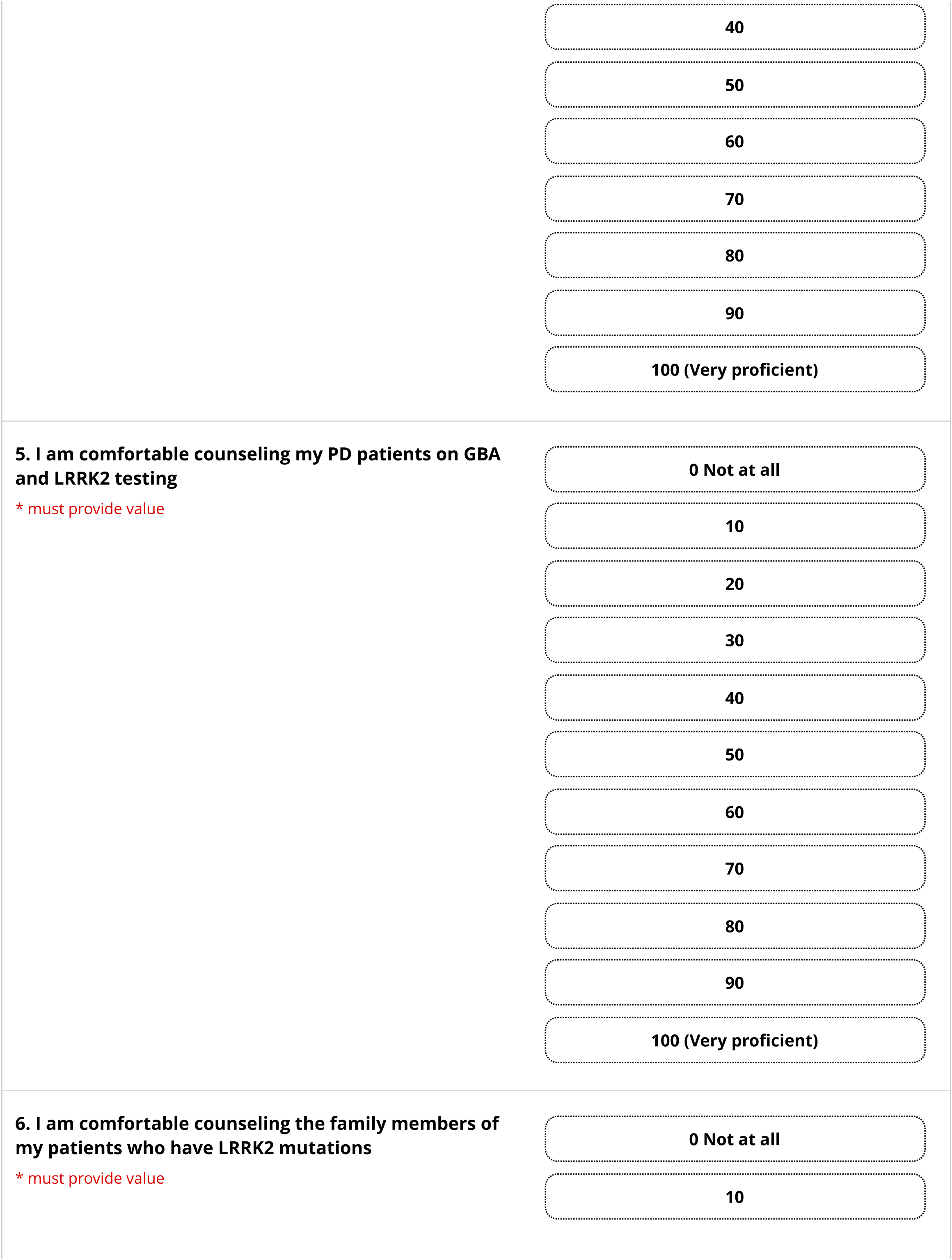
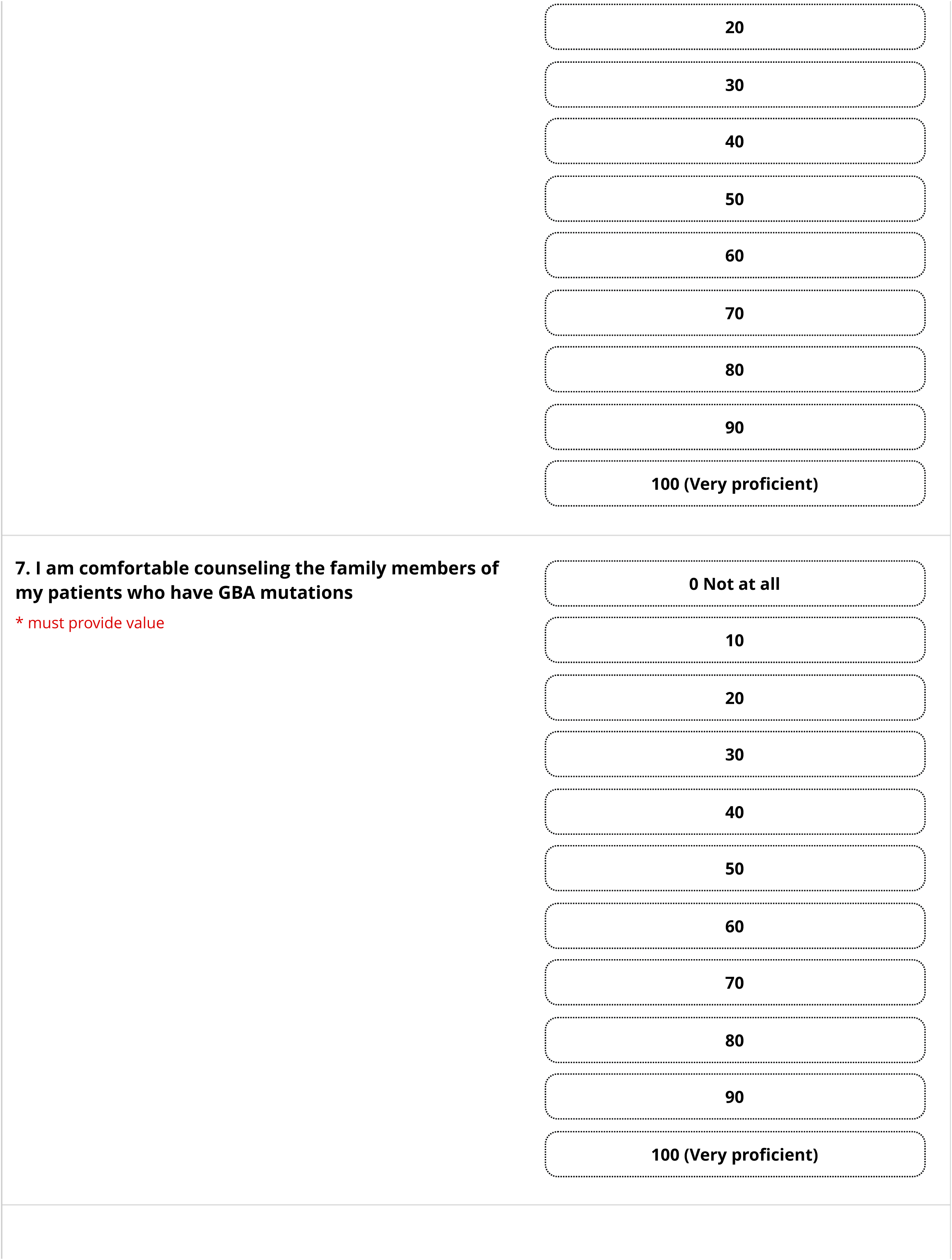
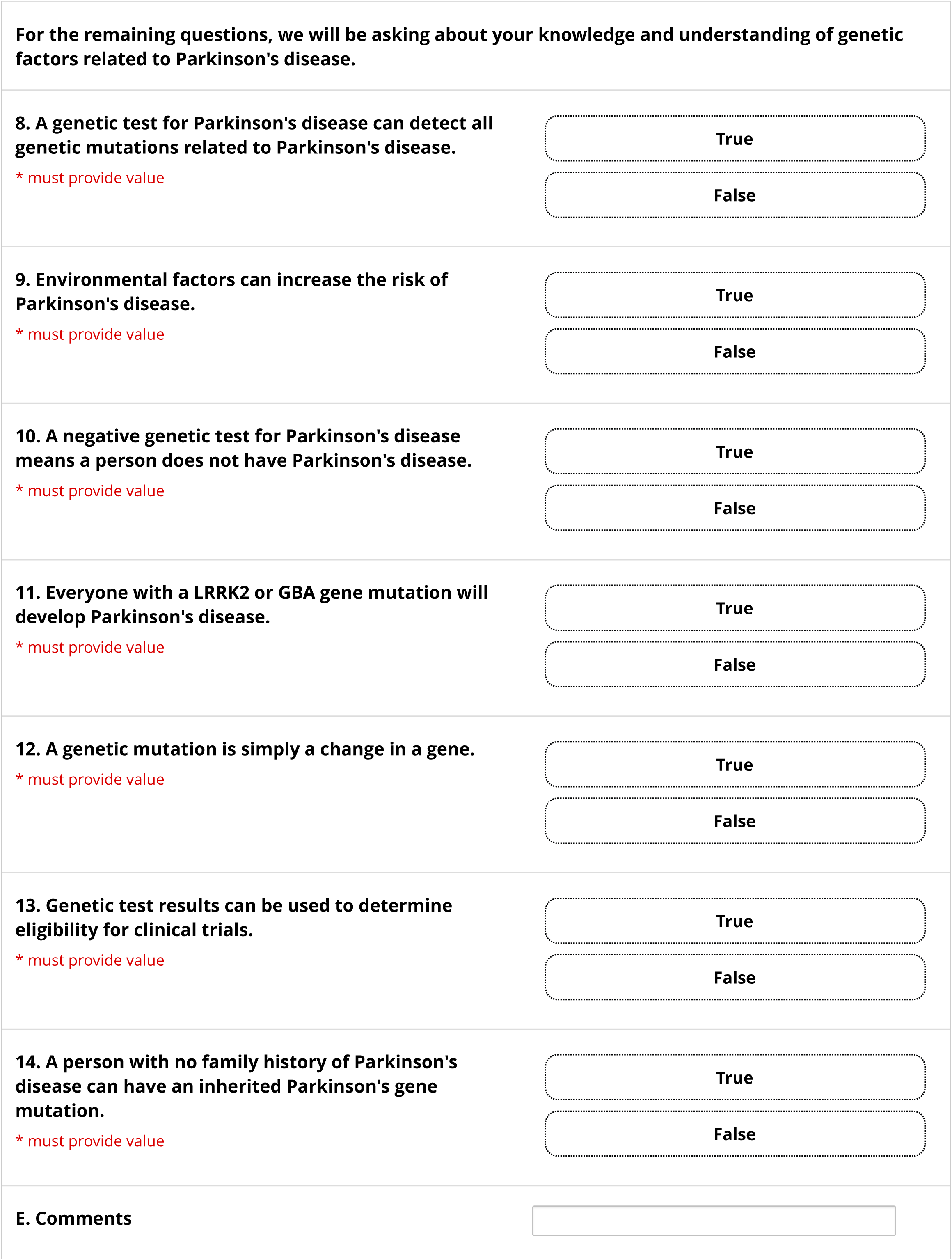
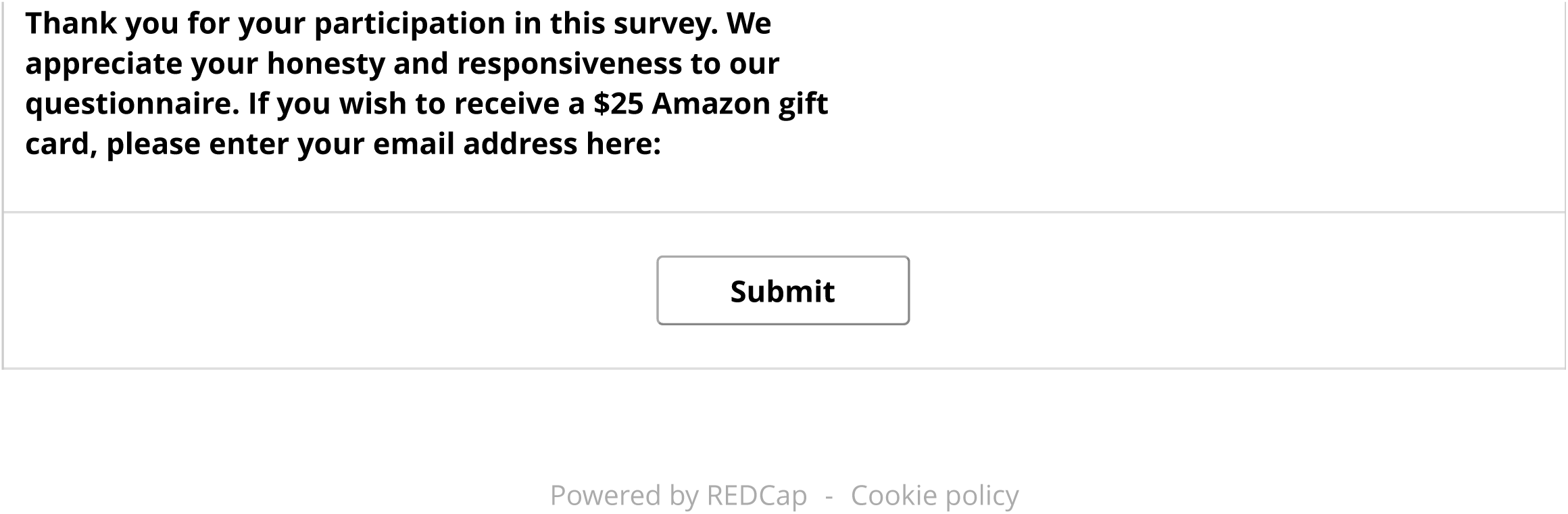

## Notes

### Competing Interest Statement

A.M.W. has received research funding from NIH/NIA, the Parkinsons Foundation, participated in clinical trials sponsored by Amylyx Pharmaceuticals, Roche/Genentech, Biogen, Ono pharmaceuticals, Bial pharmaceuticals, research funding from BioSensics, and has received consulting fees from Genentech, Ono Pharmaceuticals, and Arvinas. R.N.A. research is funded by the Michael J. Fox Foundation, the Silverstein Foundation, the Parkinsons Foundation and the Aufzien Family Center for the Prevention and Treatment of Parkinsons Disease. He received consultation fees from Bexxion, Biogen, Biohaven, Capsida, Gain Therapeutics, Genzyme/Sanofi, Janssen, SK Biopharmaceuticals, Takeda and Vanqua Bio. M.N. has received grant support for Centers of Excellence from the Parkinson Foundation, and compensation for PD GENEration Steering Committee, ACTIVATE trial sponsored by Bial, and KINECT-HD trial sponsored by Neurocrine. M.N. is Co-chair, Genetics and Environment Working group and Member, mentorship committee, Parkinson Study Group. The other authors declare no conflicts of interest.

### Funding Statement

This study did not receive any funding

### Author Declarations

IRB of Advarra reviewed and gave ethical approval for this work.

